# A Decade of CDC FluSight Influenza Forecasting

**DOI:** 10.64898/2026.06.05.26354941

**Authors:** Annabella G. Hines, Sarabeth M. Mathis, Michael A. Johansson, Carrie Reed, Matthew Biggerstaff, Rebecca K. Borchering

**Affiliations:** Influenza Division, National Center for Immunization and Respiratory Diseases, Centers for Disease Control and Prevention, United States; STI Federal, Chippewa Government Solutions, United States; Network Science Institute and Bouvé College of Health Sciences, Northeastern University, Boston, Massachusetts

## Abstract

Since the U.S. 2013/14 influenza season, the CDC’s FluSight Challenge has provided a platform for evaluating influenza forecasting models and fostering collaboration across institutions. The Challenge aims to improve the science and enhance the utility of infectious disease forecasts for public health decision making. We analyzed ten years of submitted forecasts (2014/15-2019/20 (influenza-like illness seasons) and 2021/22-2024/25 (hospital admissions seasons)) across a range of model types, including statistical, mechanistic, machine learning, and hybrid models. Influenza-like illness (ILI) forecasts were evaluated using the exponentiated logarithmic score (skill metric) while hospital admissions forecasts were evaluated using the log transformed relative Weighted Interval Score. Corresponding potential performance differences were assessed using Wilcoxon rank-sum tests, and associations with team participation history were evaluated using Spearman’s rank correlation. Model performance varied by season, and no single model type consistently outperformed others. In ILI seasons, statistical models generally performed better than mechanistic and machine learning models, though consistent differences were not observed in more recent hospital admissions seasons. Ensemble forecasts showed better overall performance across seasons, and the CDC’s FluSight ensemble ranked among the top-performing forecasts every year. We also found a positive correlation between forecast accuracy and the number of years a team participated in the Challenge, with statistically significant associations in four seasons. These findings highlight the benefits of ensemble approaches and sustained engagement in improving forecasting performance, while also underscoring the continued value of forecast evaluation before and following the COVID-19 pandemic. Insights from the FluSight Challenge can guide future infectious disease forecasting efforts and support more effective public health preparedness.

## Background

Infectious disease forecasting is a vital tool for supporting public health efforts by predicting the timing and intensity of disease activity, which helps inform communication strategies and surveillance activities [1]. In 2013, the Centers for Disease Control and Prevention (CDC) launched the Predict the Influenza Season (FluSight) Challenge to improve influenza forecast accuracy and utility for situational awareness and to help inform public health decision-making [2]. This initiative aimed to enhance reliability of predictive models while fostering cross-sector collaboration by inviting private sector, government, and academic teams to contribute forecasts. These forecasts were generated using a variety of model types, including statistical, mechanistic, hybrid, and machine learning-based approaches, each with unique strengths and challenges. This collaborative framework also facilitated evaluation utilizing pre-specified outcome data and scoring metrics, allowing for the direct comparison of forecast performance. FluSight has evolved over time to address shifting public health needs and changes in data availability [3], especially as the COVID-19 pandemic reshaped the landscape of influenza surveillance [4]. Several other forecasting collaborations and consortiums have grown from the foundation of FluSight, which pioneered the idea of a collaborative forecasting hub, including the COVID-19 Forecast Hub and European forecasting hubs, as well as Scenario Modeling Hubs [5-8].

Early seasons of the FluSight Challenge focused on predicting seasonal targets such as peak timing and intensity, as well as short-term targets forecasting influenza-like illness (ILI) activity one to four weeks into the future [2,9,10]. FluSight was paused during the 2020/21 season due to disruptions in influenza activity caused by the COVID-19 pandemic, then resumed in 2021/22 with a shift in focus to laboratory-confirmed influenza hospital admissions using data from the National Healthcare Safety Network (NHSN) [3,11]. Over time, the Challenge’s scoring methods and forecast formats have evolved to reflect changes in data and advances in modeling approaches and data infrastructure. While forecast accuracy has varied across geographic regions and seasons, each season has contributed valuable insights into improving the accuracy and utility of influenza forecasts.

In this work, we analyze the contributions and performance of forecasting models over the first 10 influenza seasons of the FluSight Challenge. We also explore whether there are common characteristics among the top-performing models [12]. We examine how the distribution of model types has evolved through time along with the changing methodology, surveillance, and data availability. Additionally, we discuss the impact of external factors such as the COVID-19 pandemic on forecasting performance and methodology, and how these developments have influenced influenza forecasting.

## Results

### Description of FluSight Challenge Characteristics Over Time

Key characteristics of each FluSight Challenge season varied from 2013/14 to 2024/25, including number of teams participating, number of models contributed, model performance, forecast targets, geographic scale, and season-specific changes (Table 1). The number of submitting models generally increased across seasons, from 13 models in 2013/14 to 38 in 2024/25. During the ILI seasons (2013/14 through 2019/20), forecast targets included onset week, peak timing, and peak intensity as reported in outpatient influenza-like illness (ILI) data, with forecasts made at both the national level and for Health and Human Services (HHS) regions [2]. In the 2014/15 season, the Challenge also introduced short-term targets predicting ILI percentages one to four weeks ahead. The 2014/15 season also introduced standardized forecast formats and proper scores to allow transparent quantitative comparison of model performance [9]. In the 2015/16 season, the CDC created a FluSight ensemble model that averaged forecasts from submitted eligible models to use in public communications and performance evaluations [10]. The Challenge has consistently emphasized real-time forecast submissions and collaboration with public health partners, including partnering with the Council of State and Territorial Epidemiologists (CSTE) beginning in 2017 to enhance operational utility [13].

**Table 1.**
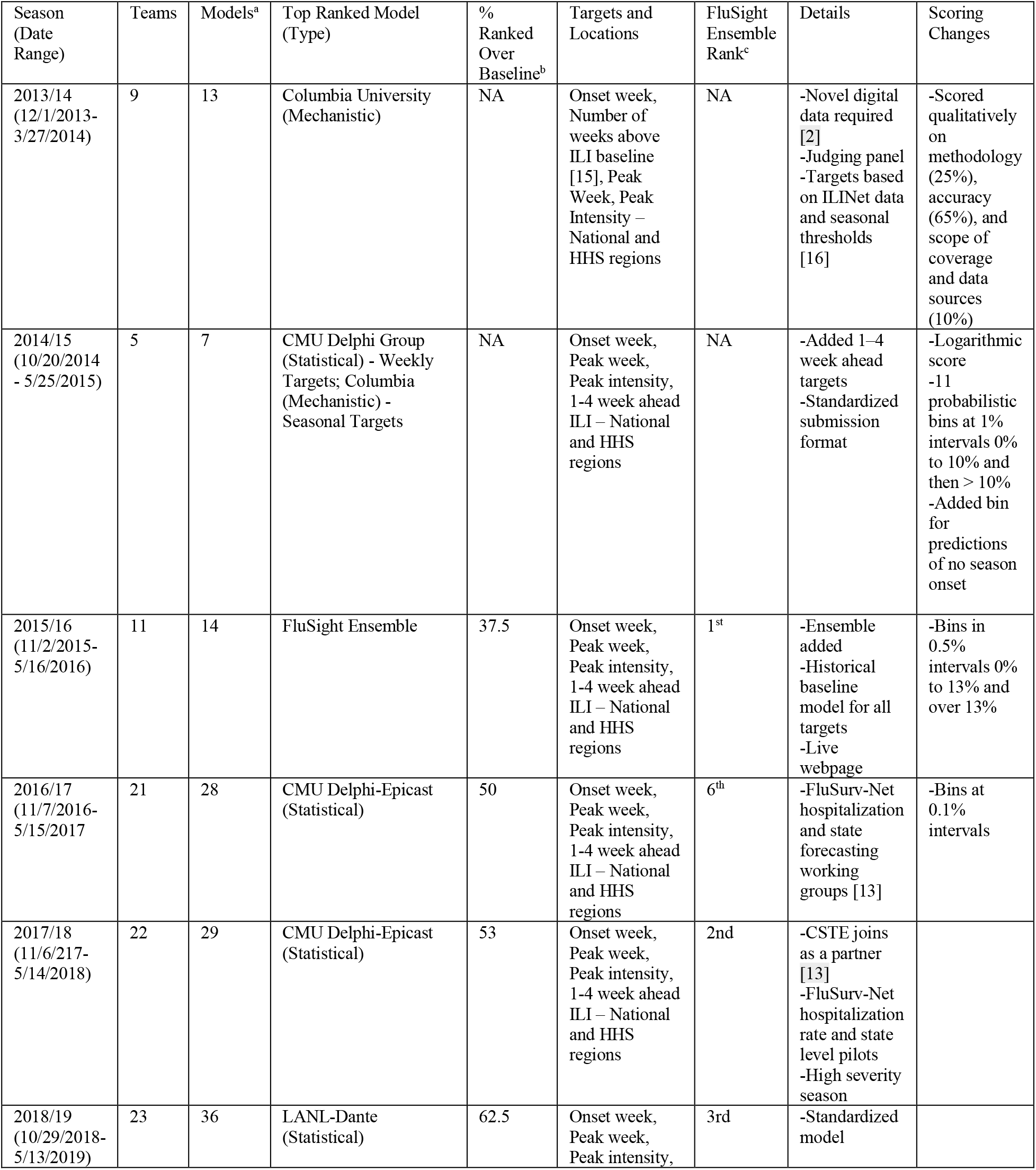
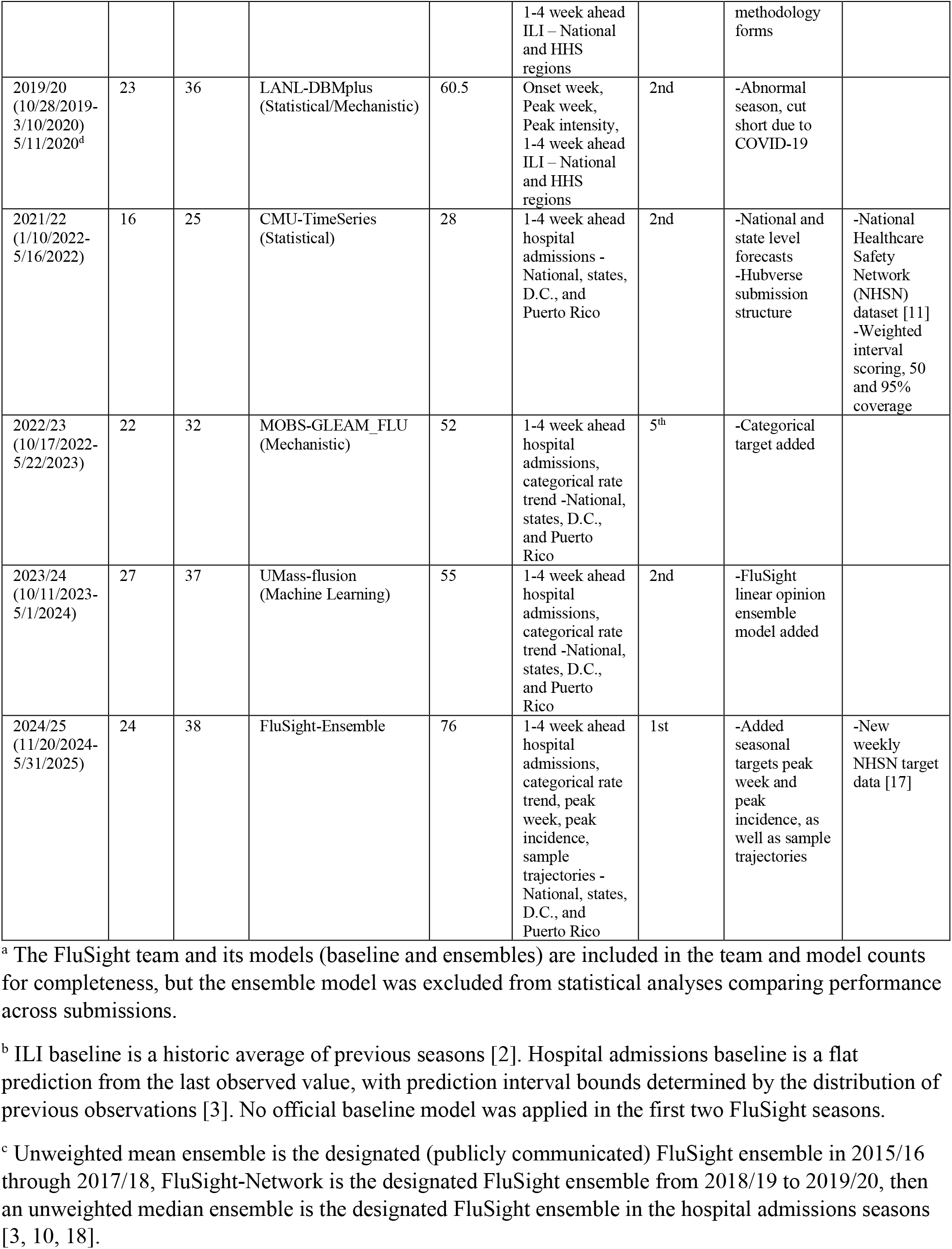
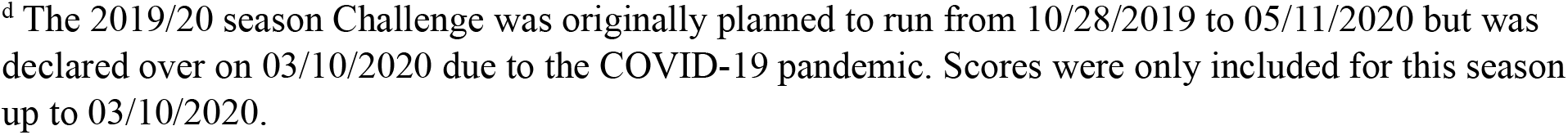
Overview of key details and guidelines for each of the 11 influenza seasons with FluSight forecasts, including team and model information, top-ranked models, and scoring changes.

| Season (Date Range) | Teams | Models<sup>a</sup> | Top Ranked Model (Type) | % Ranked Over Baseline<sup>b</sup> | Targets and Locations | FluSight Ensemble Rank<sup>c</sup> | Details | Scoring Changes |
| --- | --- | --- | --- | --- | --- | --- | --- | --- |
| 2013/14 (12/1/2013-3/27/2014) | 9 | 13 | Columbia University (Mechanistic) | NA | Onset week, Number of weeks above ILI baseline [15], Peak Week, Peak Intensity – National and HHS regions | NA | -Novel digital data required [2]<br>-Judging panel<br>-Targets based on ILINet data and seasonal thresholds [16] | -Scored qualitatively on methodology (25%), accuracy (65%), and scope of coverage and data sources (10%) |
| 2014/15 (10/20/2014 - 5/25/2015) | 5 | 7 | CMU Delphi Group (Statistical) - Weekly Targets; Columbia (Mechanistic) - Seasonal Targets | NA | Onset week, Peak week, Peak intensity, 1-4 week ahead ILI – National and HHS regions | NA | -Added 1–4 week ahead targets<br>-Standardized submission format | -Logarithmic score<br>-11 probabilistic bins at 1% intervals 0% to 10% and then > 10%<br>-Added bin for predictions of no season onset |
| 2015/16 (11/2/2015-5/16/2016) | 11 | 14 | FluSight Ensemble | 37.5 | Onset week, Peak week, Peak intensity, 1-4 week ahead ILI – National and HHS regions | 1<sup>st</sup> | -Ensemble added<br>-Historical baseline model for all targets<br>-Live webpage | -Bins in 0.5% intervals 0% to 13% and over 13% |
| 2016/17 (11/7/2016-5/15/2017) | 21 | 28 | CMU Delphi-Epicast (Statistical) | 50 | Onset week, Peak week, Peak intensity, 1-4 week ahead ILI – National and HHS regions | 6<sup>th</sup> | -FluSurv-Net hospitalization and state forecasting working groups [13] | -Bins at 0.1% intervals |
| 2017/18 (11/6/2017-5/14/2018) | 22 | 29 | CMU Delphi-Epicast (Statistical) | 53 | Onset week, Peak week, Peak intensity, 1-4 week ahead ILI – National and HHS regions | 2<sup>nd</sup> | -CSTE joins as a partner [13]<br>-FluSurv-Net hospitalization rate and state level pilots<br>-High severity season |  |
| 2018/19 (10/29/2018-5/13/2019) | 23 | 36 | LANL-Dante (Statistical) | 62.5 | Onset week, Peak week, Peak intensity, | 3<sup>rd</sup> | -Standardized model |  |
|  |  |  |  |  | 1-4 week ahead<br>ILI – National<br>and HHS<br>regions |  | methodology<br>forms |  |
| 2019/20<br>(10/28/2019-3/10/2020)<br>5/11/2020<sup>d</sup> | 23 | 36 | LANL-DBMplus<br>(Statistical/Mechanistic) | 60.5 | Onset week,<br>Peak week,<br>Peak intensity,<br>1-4 week ahead<br>ILI – National<br>and HHS<br>regions | 2nd | -Abnormal<br>season, cut<br>short due to<br>COVID-19 |  |
| 2021/22<br>(1/10/2022-5/16/2022) | 16 | 25 | CMU-TimeSeries<br>(Statistical) | 28 | 1-4 week ahead<br>hospital<br>admissions -<br>National, states,<br>D.C., and<br>Puerto Rico | 2nd | -National and<br>state level<br>forecasts<br>-Hubverse<br>submission<br>structure | -National<br>Healthcare<br>Safety<br>Network<br>(NHSN)<br>dataset [11]<br>-Weighted<br>interval<br>scoring, 50<br>and 95%<br>coverage |
| 2022/23<br>(10/17/2022-5/22/2023) | 22 | 32 | MOBS-GLEAM_FLU<br>(Mechanistic) | 52 | 1-4 week ahead<br>hospital<br>admissions,<br>categorical rate<br>trend -National,<br>states, D.C.,<br>and Puerto<br>Rico | 5<sup>th</sup> | -Categorical<br>target added |  |
| 2023/24<br>(10/11/2023-5/1/2024) | 27 | 37 | UMass-flusion<br>(Machine Learning) | 55 | 1-4 week ahead<br>hospital<br>admissions,<br>categorical rate<br>trend -National,<br>states, D.C.,<br>and Puerto<br>Rico | 2nd | -FluSight<br>linear opinion<br>ensemble<br>model added |  |
| 2024/25<br>(11/20/2024-5/31/2025) | 24 | 38 | FluSight-Ensemble | 76 | 1-4 week ahead<br>hospital<br>admissions,<br>categorical rate<br>trend, peak<br>week, peak<br>incidence,<br>sample<br>trajectories -<br>National, states,<br>D.C., and<br>Puerto Rico | 1st | -Added<br>seasonal<br>targets peak<br>week and<br>peak<br>incidence, as<br>well as sample<br>trajectories | -New<br>weekly<br>NHSN target<br>data [17] |

<sup>a</sup> The FluSight team and its models (baseline and ensembles) are included in the team and model counts for completeness, but the ensemble model was excluded from statistical analyses comparing performance across submissions.
<sup>b</sup> ILI baseline is a historic average of previous seasons [2]. Hospital admissions baseline is a flat prediction from the last observed value, with prediction interval bounds determined by the distribution of previous observations [3]. No official baseline model was applied in the first two FluSight seasons.
<sup>c</sup> Unweighted mean ensemble is the designated (publicly communicated) FluSight ensemble in 2015/16 through 2017/18, FluSight-Network is the designated FluSight ensemble from 2018/19 to 2019/20, then an unweighted median ensemble is the designated FluSight ensemble in the hospital admissions seasons [3, 10, 18].
<sup>d</sup> The 2019/20 season Challenge was originally planned to run from 10/28/2019 to 05/11/2020 but was declared over on 03/10/2020 due to the COVID-19 pandemic. Scores were only included for this season up to 03/10/2020.

During the hospital admission period (2021/22 onward), models predicted laboratory-confirmed influenza hospital admissions nationally and for all states, Washington D.C., and Puerto Rico using a surveillance system that became available during the pandemic [11]. The 2021/22 season marked a shift to quantile-based forecast submissions, the adoption of the weighted interval score (WIS), and regular evaluation of prediction interval coverage (e.g., 50% and 95%) to provide a more comprehensive evaluation of forecast performance [3, 14]. The 2021/22 FluSight season officially implemented Hubverse standards, which provided a more flexible standardized structure for organizing and validating forecast submissions, improved submission consistency and enabled broad use of shared tools and infrastructure [5]. In the 2022/23 season, FluSight added new categorical rate trend targets (e.g. probability of increase/decrease in influenza hospital admissions) [3]. Seasonal targets including peak week and peak incidence were brought back for the 2024/25 season.

### Performance by Model Type

The number of teams participating and models submitted to the FluSight Challenge generally increased over time, with a temporary decrease in participation during the first hospital admissions season. While mechanistic and statistical models dominated submissions across all seasons, the proportion of machine learning models increased in later years from 17% of submitted models in ILI seasons to 29% in hospital admissions seasons Beginning in 2015/16, each season featured one designated FluSight ensemble forecast, occasionally accompanied by different pilot ensemble forecasts (Table 1). Overall, statistical models made up the largest share of submissions (41%), followed by machine learning (24%), mechanistic (21%), statistical/mechanistic combinations (9%), and FluSight ensembles (5%). The uneven distribution of model types may have influenced the performance results reported in later sections (Figure 1).

**Figure 1.**
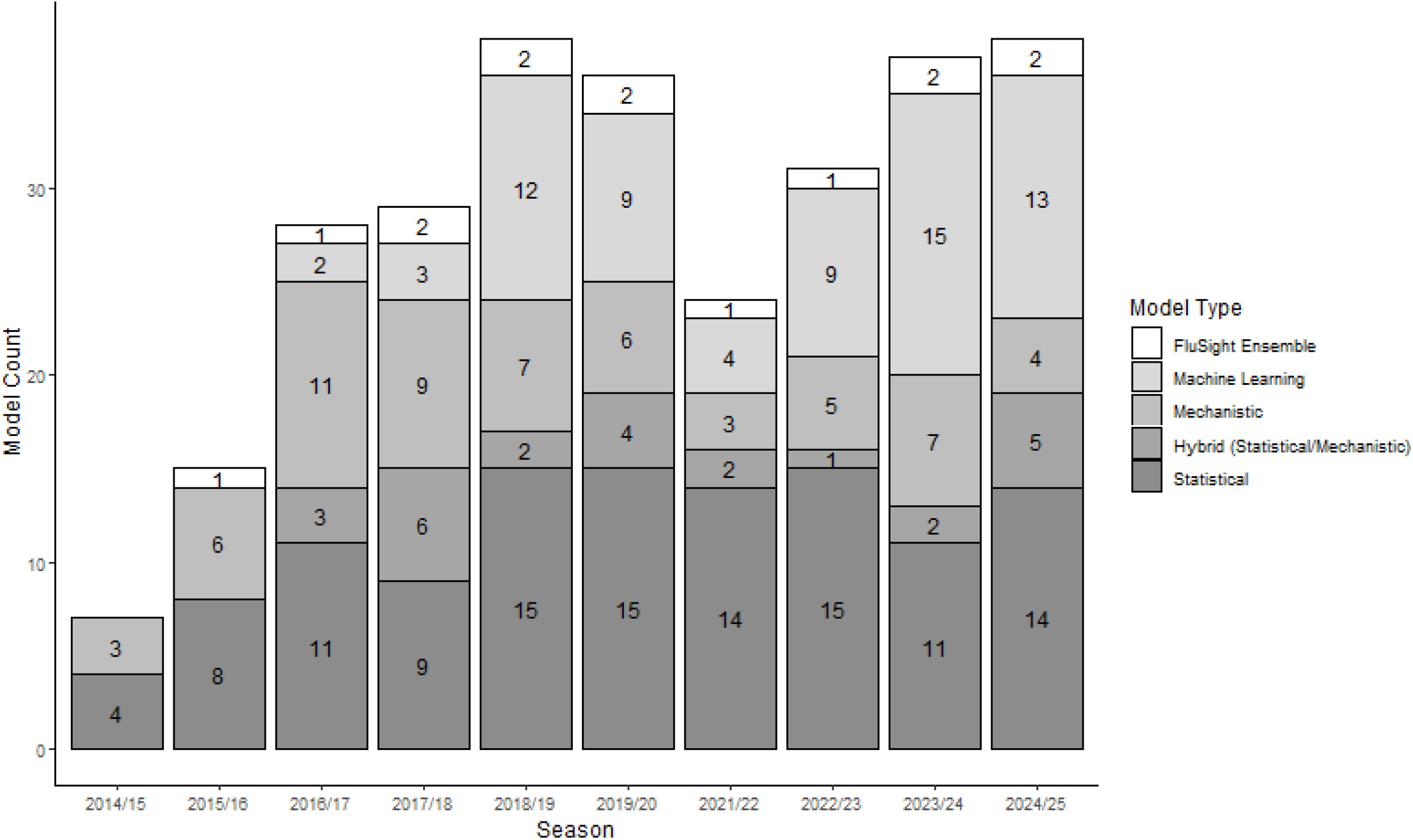
Distribution of model types from the 2014/15 season to the 2024/25 season.

Since different scoring methods were used before and after the emergence of COVID-19, we assessed model performance separately for the ILI seasons (2014/15-2019/20) and the hospital admissions seasons (2021/22-2024/25). We compared the distribution of performance scores across four submitted model categories: statistical, machine learning, mechanistic, and statistical/mechanistic hybrids (models incorporating elements of both). Pairwise comparisons across combined ILI seasons indicated that statistical and statistical/mechanistic hybrid models outperformed mechanistic models (p < 0.05). At the individual season level, statistical models performed significantly better than mechanistic models in 2018/19, and better than machine learning models in 2019/20 (p < 0.05) (Figure 2). No significant performance differences were observed between model types in other ILI seasons (see Supplementary Table 2).

**Figure 2.**
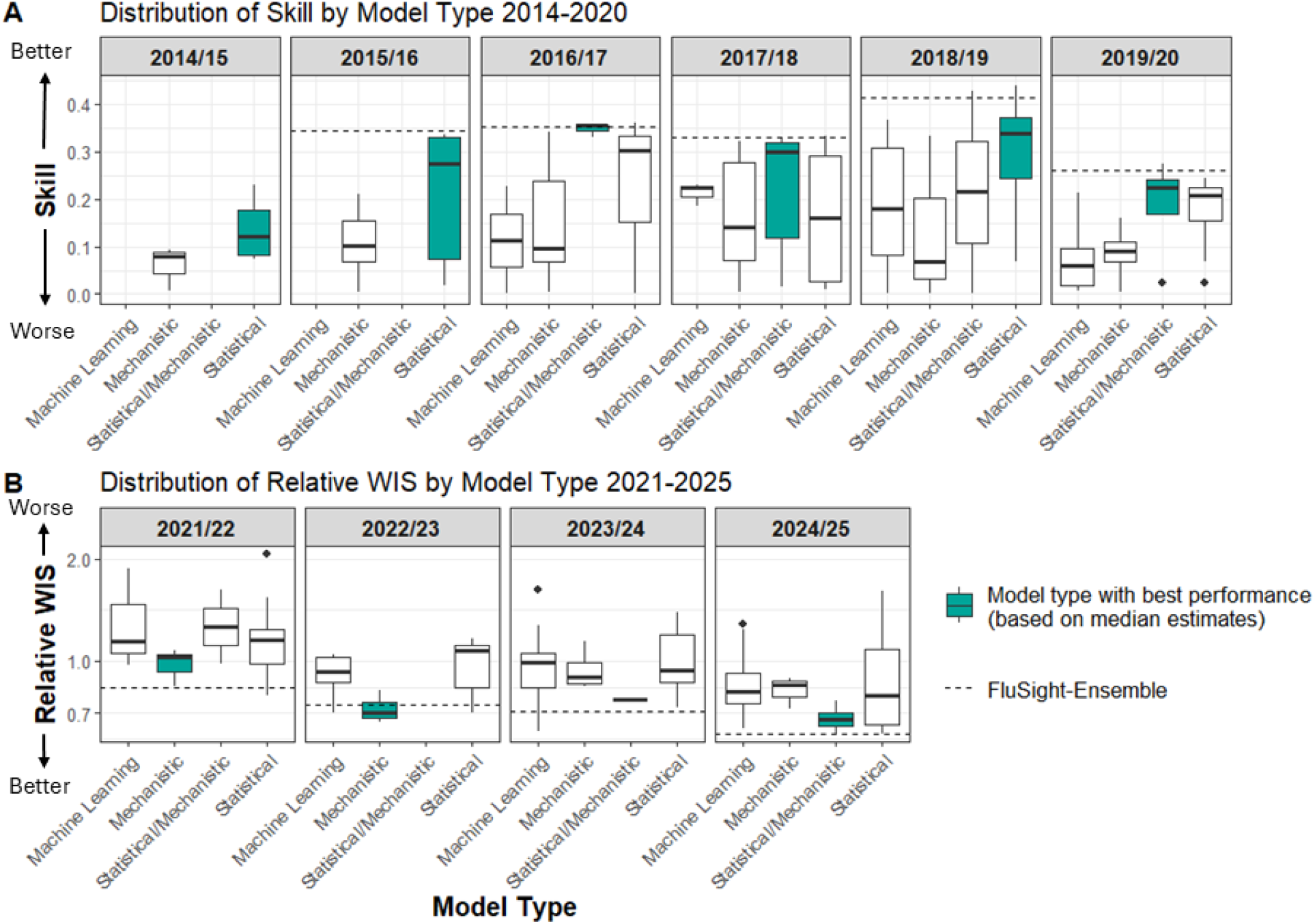
Boxplots comparing model performance by model type across influenza seasons. **A**. Influenza-like Illness seasons (2014/15-2019/20): Distribution of forecast skill by model type; higher values indicate better performance. **B**. Hospital admissions seasons (2021/22-2024/25): Distribution of relative Weighted Interval Scores (WIS) by model type; lower values indicate better performance. Relative WIS is displayed on a logarithmic scale with baseline performance equal to one. FluSight ensemble model scores are represented as a dashed line and included as a visual benchmark, but were excluded from the statistical analysis. The model type highlighted in teal each season is the category with the best median score.

For influenza hospitalization seasons, no significant differences in model performance were observed within individual seasons or pooled seasons (Supplementary Table 2). Across the 11 seasons, statistical models ranked first in five seasons, mechanistic models in three, FluSight ensembles in two, and machine learning and statistical/mechanistic hybrid models each topped the rankings once. In the 2014/15 season, seasonal and short-term targets were scored separately, with a mechanistic model ranked first for seasonal targets and a statistical model ranked first for short-term targets. A full list of the top five ranked models by season, including their model types, is provided in Supplementary Table 3.

### Performance by Ensemble Status

Submitted models were categorized as “ensemble” or “single” based on their methodology provided in the associated metadata, as described in the methods. Across all included seasons, 38% of submitted models were categorized as ensembles, with seasonal proportions ranging from 34% to 48%. Submitted forecasts which used ensembles were significantly better than single models when pooled across ILI seasons (p<0.001) (Supplementary Figure 2). When analyzing individual seasons, ensemble models outperformed single models in terms of the forecast skill metric in most ILI seasons (p < 0.05), except for the 2014/15 and 2015/16 seasons (Figure 3A). Although no hospital admissions seasons showed statistically significant differences, ensemble models generally exhibited higher median skill in the ILI period and lower relative WIS in the hospital admissions period, both indicating better performance (Figure 3).

**Figure 3.**
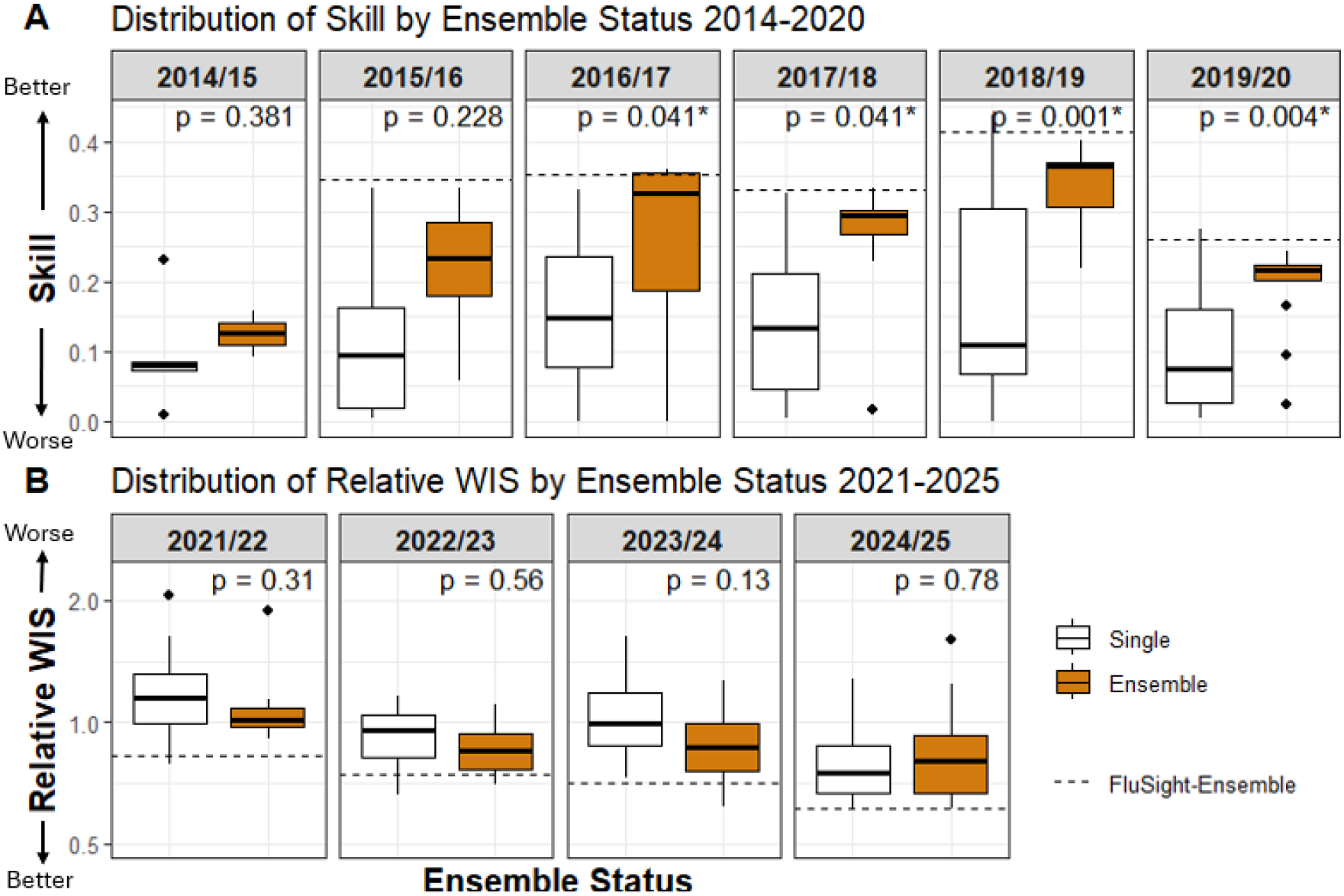
Boxplots comparing submitted ensemble and single model performance across influenza seasons. **A**. Influenza-like Illness seasons (2014/15-2019/20): Distribution of skill scores by ensemble or single model status; higher values indicate better performance. **B**. Hospital admissions seasons (2021/22-2024/25): Distribution of relative Weighted Interval Scores (WIS) by ensemble or single model status; lower values indicate better performance. Asterisks (*) denote statistically significant differences between groups (*p* < 0.05). Relative WIS is displayed on a logarithmic scale with baseline performance equal to one. Dashed lines indicate the FluSight-ensemble performance for each season.

FluSight ensemble models (FluSight-Network, FluSight-ensemble) were excluded from the model-type analysis since they are a combination of all eligible models submitted to FluSight. The FluSight baseline model was also excluded from this analysis as it is not considered a submitted model. However, FluSight ensembles consistently performed well (Figure 3), ranked highly every season, and weres in the top five forecasts for eight out of the nine seasons in which a FluSight ensemble was produced (Supplemental Table 3). This included high severity U.S. influenza seasons (e.g., 2017/18) and influenza seasons with atypical timing (e.g., 2024/25) (Supplementary Table 1).

### Performance by Years of Participation

To explore the relationship between years of prior participation in FluSight and model performance, we examined the number of seasons a team had previously submitted forecasts. The analysis revealed that in all seasons, more years of FluSight participation were positively correlated with improved performance (higher skill in ILI seasons and lower relative WIS in hospital admissions seasons), with statistically significant correlations (p < 0.05) observed in the 2016/17, 2017/18, 2018/19, 2019/20, 2022/23, and 2024/25 seasons (Figure 4), as well as when aggregating all hospital admissions seasons and all ILI seasons(See Supplementary Figure 3). It is important to note that the participation years reflect the team’s accumulated FluSight experience, rather than the number of times a particular model has been submitted to the Challenge. While teams often participated in multiple years, the models themselves frequently changed from season to season. There was a high return rate for forecasting teams, with between 50 and 100% of teams returning each season from the previous one. This approach accounts for the overall FluSight experience of the team, which could include the development of new models that may improve upon previous ones. However, prior participation did not universally translate to improved performance as there were some experienced teams that performed poorly in some seasons, suggesting other factors also play a role.

**Figure 4.**
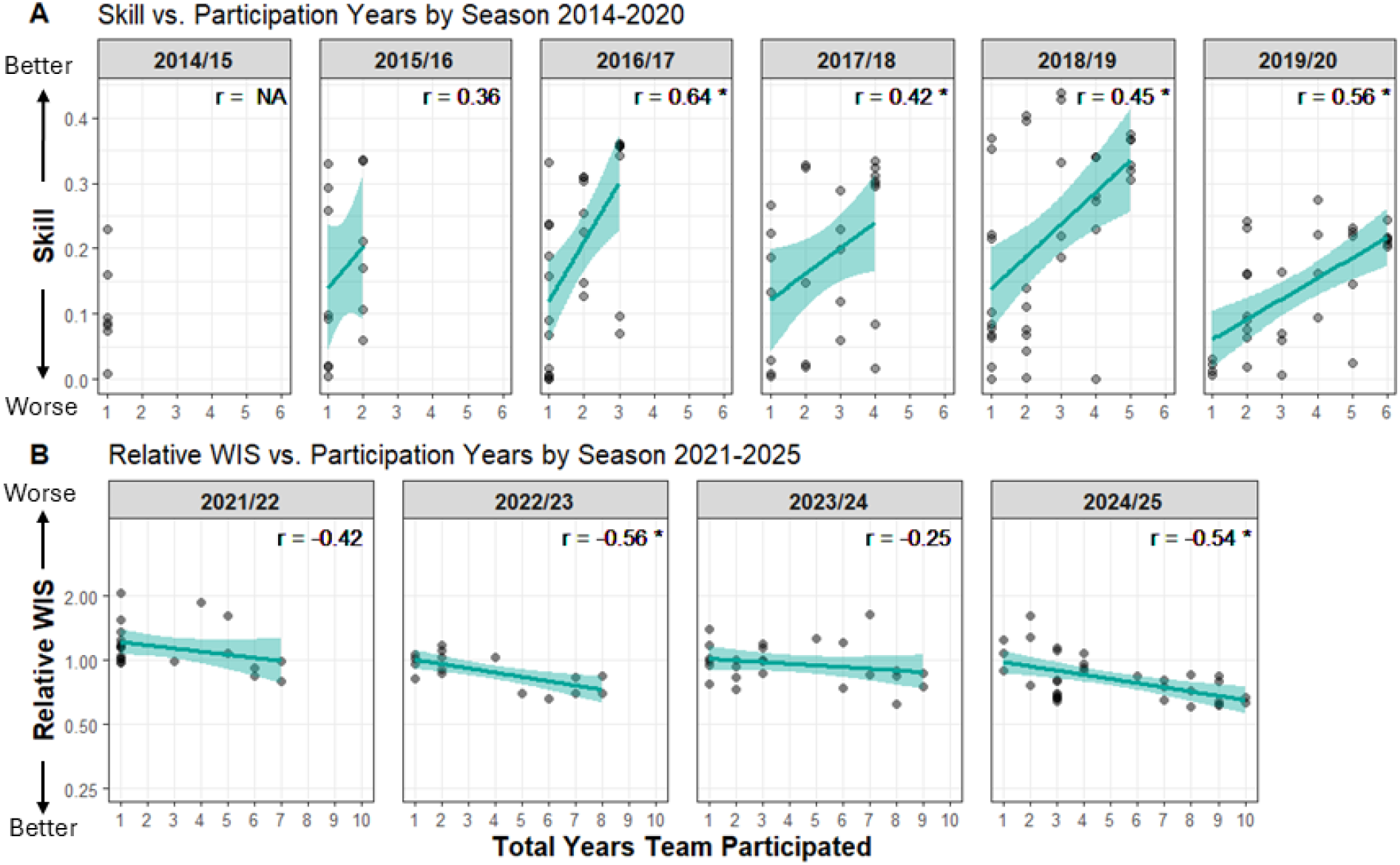
Scatter plots showing the relationship between model performance and number of participation years across influenza seasons. Solid lines and shaded bands show fitted trends and 95% confidence intervals (see Methods for details). Spearman correlation coefficients are shown for each season. **A**. Influenza-like Illness seasons (2014/15-2019/20): Forecast skill plotted against years of participation. Higher skill values indicate better performance. **B**. Hospital admissions seasons (2021/22-2024/25): Relative Weighted Interval Score (WIS) plotted against years of participation. Lower relative WIS values indicate better performance. Relative WIS is displayed on a logarithmic scale with baseline performance equal to one. Asterisks (*) indicate statistically significant correlations (*p* < 0.05).

To assess the impact of prior (ILI period) FluSight participation on model performance in the hospital admissions seasons, we divided the hospital admissions model submissions into two groups: those submitted by teams that had participated in FluSight both periods, and those submitted by teams that participated only in the hospital admissions period (2021/22 or later). Across all hospital admissions seasons (2021/22-2024/25), 25 of the 53 models (47%) submitted from teams that never submitted ILI forecasts scored better than the baseline forecast (i.e., had a relative WIS less than 1). In contrast, 34 of the 43 model submissions (79%) from teams with experience in ILI forecasting outperformed the baseline (Figure 5). A Wilcoxon rank sum test revealed a significant difference between the two groups (p = 0.0003), indicating that teams with prior participation tended to perform better in the hospital admissions seasons than teams without prior FluSight experience.

**Figure 5.**
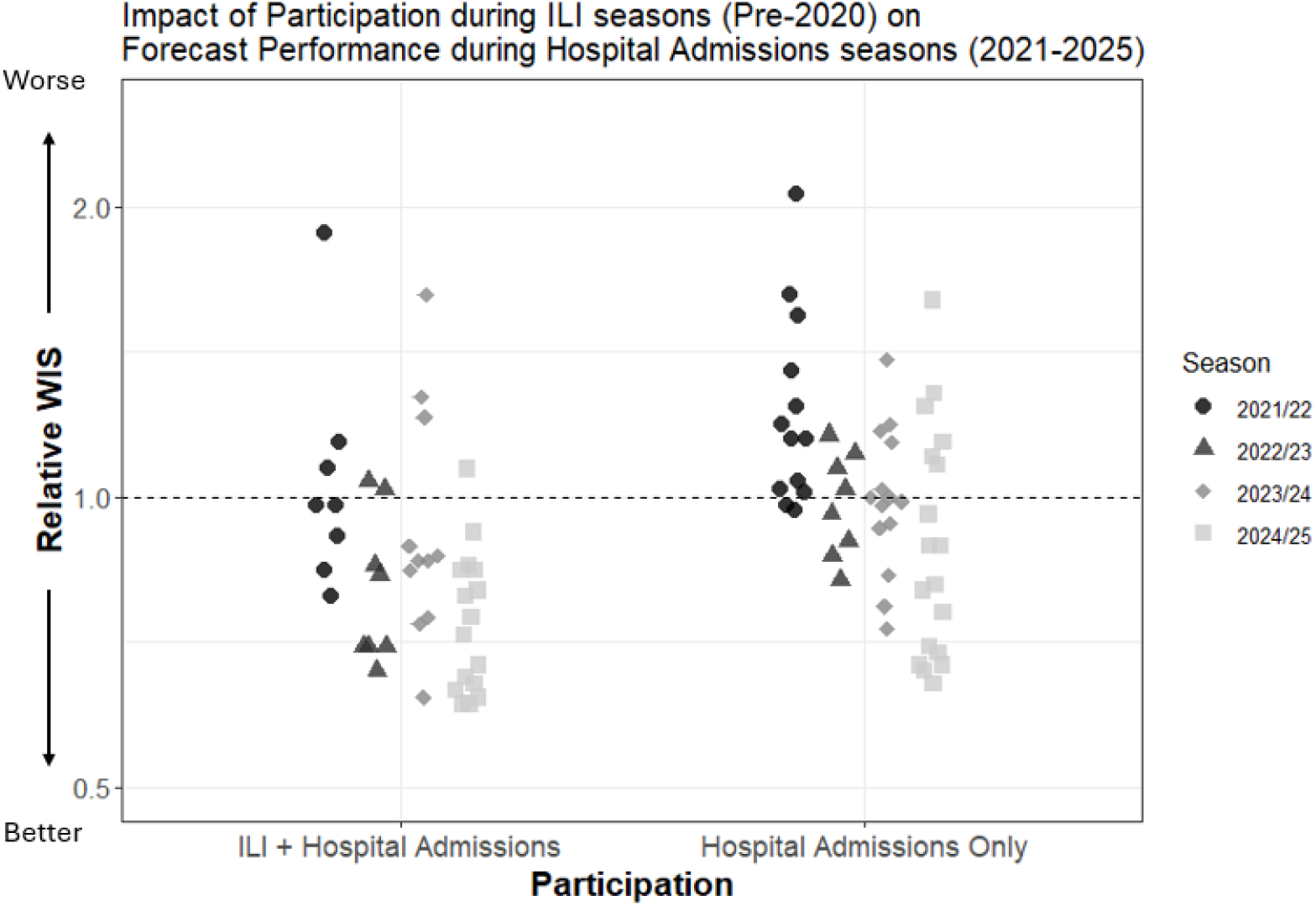
Scatter plot showing the relative Weighted Interval Score (rWIS) of each model during hospital admissions seasons, grouped by participation history (ILI and hospital admissions seasons vs. hospital admissions seasons only). The horizontal line at relative WIS = 1 indicates the baseline model performance. Models with relative WIS < 1 outperformed the baseline; lower values indicate better performance. Relative WIS is displayed on a logarithmic scale.

## Discussion

The FluSight Challenge has been conducted consistently for over a decade, and as of May 4th, 2026 continues, creating a unique repository of influenza forecasts that offers insight into the evolution of infectious disease forecasting [1, 19]. The duration of the Challenge, combined with high team retention from season to season, provides a comprehensive overview of the forecasting landscape over time, revealing key patterns and trends. By fostering a platform for the systematic comparison of influenza forecasts across multiple seasons, FluSight has enabled researchers and public health professionals to track how forecasting techniques evolve in response to changes in the disease landscape, data availability, and modeling approaches. At the same time, it has supported improvements in how forecasts are developed and evaluated and has revealed forecasting methods of increasing utility to public health decision-making.

A primary finding of this analysis is that ensemble models consistently outperformed individual models even in high-severity or unusually timed influenza seasons, reinforcing the value of combining multiple forecasting approaches [3, 9, 10]. Along with ensembles in general, the FluSight ensembles performed well every season, specifically ranking in the top five models in eight out of the nine seasons since it was first included (Supplemental Table 3). This finding has been supported by results in other forecasting challenges and contexts, like the COVID-19 forecast hub and Dengue forecasting efforts [6, 20]. Having shown to be generally high performing, ensemble methods can provide a robust choice for forecasting, especially when individual contributing model performance varies across seasons [5,21,22]. This is crucial in the context of infectious disease forecasting, where high variability and unpredictable epidemiological patterns can challenge individual models and communication and public health decisions must be made a priori to forecast evaluation. Ensemble models may outperform individual models by accounting for multiple forecasts, thereby increasing the robustness and reliability of predictions.

Our analysis of over a decade of forecasting results also highlights that no single model type was consistently the best performing across all seasons. Seasonal variations, such as differences in influenza season intensity or the emergence of external factors like the COVID-19 pandemic, could influence which type of model is most robust for a given season. In ILI seasons, statistical and statistical/mechanistic combination models as a group generally outperformed mechanistic models at predicting ILINet percentages (Supplementary Table 2). Pooled results for ILI seasons should be interpreted with caution, in part because scoring criteria varied, which could introduce bias. Statistical models are well-suited for capturing trends and seasonality in percentage-based metrics, particularly since they were able to be trained on extensive historical ILINet data which extends back to 1997 [23]. Conversely, in the hospital admissions period, the shift to a new dataset with little to no historic data for training, the NHSN hospital admission counts [11], may have limited the performance of statistical models [24]. Some models used an extended time series of hospital admissions scaled using available ILINet data to supplement the NHSN data for training, with the top ranked model in the 2023/24 season utilizing this approach [25]. It is important to note, however, the limited sample size for each model type per season, as well as the complexities in comparing seasons due to scoring changes and season-specific epidemiological dynamics. For example, the 2021/22 and 2022/23 influenza seasons exhibited atypical timing, which likely also increased the difficulty of forecasting. This may have particularly been a challenge for statistical models which rely heavily on previously observed data for anticipating changes in trends. There was a notable increase in the number of machine learning models over time, particularly in the hospital admissions period.

As expected, teams that participated in multiple seasons of the FluSight Challenge generally demonstrated better performance than those with fewer years of participation (Figure 4). This pattern may be attributed to the accumulation of experience and the refinement of modeling techniques to adjust to influenza forecasting challenges. Teams with a history of participation may be more able to adapt their models based on lessons learned from prior seasons, and this process of learning from past forecasting seasons may enable teams to fine-tune their models and improve their performance. This was particularly true for forecasts in the ILI period (Figure 4A) where the forecast target, scoring criteria, and seasonality of influenza dynamics were more consistent across seasons. However, we were not able to evaluate season-to-season changes in model performance associated directly with updated model methodology due to the variability in influenza seasons and data availability which may inherently change the difficulty of producing accurate forecasts. Changing forecast requirements and scoring practices also complicates directly comparing model performances across seasons. Even in the context of substantial shifts in the influenza forecasting landscape, general participation in FluSight before COVID-19 seems to have a positive impact on forecast performance after the COVID-19 pandemic (Figure 5). At the same time, systematic FluSight forecasting challenges built capacity for collaborative forecasting, enabling the timely standup of the COVID-19 Forecasting hub as well as other forecasting hubs [5-7].

There remains substantial room to characterize when particular forecasting models perform best, not only across different seasons but also within phases of a season, such as the increasing, peak, and declining phases[26]. By analyzing data over many seasons, researchers can identify which models excel under specific conditions, such as low versus high severity seasons or seasons with early versus late peaks[16]. This could also help reveal which factors, such as the dominant influenza strain, regional differences, or public health interventions, most influence model performance (Supplemental Table 1). These insights are critical for refining forecasting models and improving their utility in public health planning. Moreover, the transition of the FluSight forecast target during the COVID-19 pandemic underscores the importance of adaptability in forecasting models. Moving forward as the public health data landscape continues to change, the ability of forecasting approaches to adapt to new data sources will be crucial for maintaining and improving forecast accuracy.

In summary, the FluSight Challenge has provided invaluable advances and insights into infectious disease forecasting and collaboration over the past decade. The consistent performance of ensemble models over single models has cemented their use in applied infectious disease forecasting. Additionally, the observed improvement in forecasting accuracy among teams with extensive participation highlights the importance of maintaining forecasting systems that build experience and the ability to learn and refine modeling techniques. Over the decade of the FluSight Challenge, no single model type emerged as the most accurate approach across all seasons, and this analysis emphasized that understanding seasonal variations and contextual factors was crucial for evaluating forecasts. The shift in the FluSight forecasting target due to the availability of new data illustrates how shifts in available information could influence model performance. As public health professionals increasingly utilize forecasts for situational awareness and decision-making, ongoing efforts to enhance model accuracy through collaboration and innovation will be essential. Ultimately, insights gained from the ongoing FluSight Challenge can continue to inform future research and improve public health responses to influenza epidemics and pandemics, ensuring that forecasting remains a vital tool in managing infectious diseases.

## Methods

### Data Sources

We analyzed forecast data from the FluSight Challenge, with ILI forecasts for the 2014/15 through 2019/20 seasons and laboratory-confirmed influenza hospital admissions forecasts for the 2021/22 through 2024/25 seasons in the United States. Data were aggregated from archived repositories for previous seasons, with performance scores calculated according to each season’s specific guidelines. In the ILI seasons forecast targets included onset week, peak timing, and peak intensity as reported in outpatient influenza-like illness (ILI) data, with forecasts made at both the national level and for Health and Human Services (HHS) regions [2]. In the hospital admissions seasons, forecast targets included laboratory-confirmed hospital admissions at nationally, at the state-level, and for Washington D.C. and Puerto Rico as reported in NHSN [17]. In the hospital admissions seasons, models that submitted less than 75% of total forecasts throughout a season were excluded from analysis. We also compiled season-specific epidemiological characteristics such as severity, timing, and other relevant factors (see Supplementary Table 1) to provide additional context for interpreting model performance across seasons. The initial 2013/14 season was excluded from performance analysis due to substantial differences in forecast submission, format, and scoring methodology [2,9].

### Performance Metrics

Performance was assessed using two different proper scoring metrics corresponding to the ILI and hospital admission periods. For the ILI seasons (2014/15-2019/20), the exponentiated logarithmic score (also referred to as the skill metric) was used as the primary performance measure [10]. This score quantifies the probability assigned to the observed outcome by the forecast distribution, with exponentiation applied to make scores more interpretable across forecasts. Each forecast target (e.g., peak week, 1-week ahead) was scored per location and submission week against the observed U.S. Outpatient Influenza-like Illness Surveillance Network (ILINet) ILI value and then averaged across time and locations to produce an overall score [23]. Scoring during these seasons followed FluSight guidelines released specifically for each season, which included differences (some substantial) across years, such as variation in bin widths defining the accuracy thresholds (Table 1). The final score incorporated both short-term and seasonal target scores.

For the hospital admissions seasons (2021/22-2024/25), the relative Weighted Interval Score (rWIS), computed using log transformed admissions values (i.e. log(admissions + 1)), was employed to score forecasts of laboratory confirmed influenza hospital admissions. WIS is a robust metric for evaluating forecast accuracy, penalizing forecasts with wider, less sharp prediction intervals [3]. Forecast values and observed values were log transformed prior to calculating the WIS to normalize scores across locations, preventing larger states from disproportionately influencing the scoring [3, 27-29]. Here we analyze the scaled relative WIS, where each model’s relative skill is calculated as the geometric mean of pairwise WIS ratios across all comparator models and then scaled relative to the FluSight baseline model. Because scaled relative WIS is a ratio-based metric centered at 1, values equally distant from 1 on a multiplicative scale (e.g., 0.5 and 2) represent equivalent magnitudes of improvement and degradation relative to the baseline model. The FluSight hospital admissions baseline model predicts a flat trend from the most recent observed value each week of the influenza season and adds noise to generate the confidence interval based on the distribution of recent observations [3]. This relative comparison allows the assessment of model performance against a baseline forecast which is also subject to forecasting challenges such as changing trends. The use of log-transformed values and relative scoring metrics also helps reduce the extent to which locations with larger hospitalization counts disproportionately influence aggregate performance measures.

### Model Types

Models were classified into one of five categories based on their underlying methodologies, which were either explicitly stated or inferred from manual inspection of model metadata:

- Mechanistic Models (MECH): Models that incorporate compartmental modelling, e.g., Susceptible-Exposed-Infectious-Recovered (SEIR) models.
- Statistical Models (STAT): Models that rely on statistical techniques such as time-series analysis and generalized linear models.
- Machine Learning Models (ML): Models that incorporate machine learning components, such as random forests, neural networks, or support vector machines, including models that would otherwise be categorized as mechanistic or statistical.
- Statistical/Mechanistic Hybrid Models (STAT/MECH): Models that combine elements from both statistical and mechanistic approaches.
- FluSight Ensemble: Forecast generated from a combination of submitted models. From the 2015/16 to 2017/18 seasons, an unweighted mean ensemble was used. For 2018/19 and 2019/20 season, the FluSight Network ensemble weighted on past model performance was utilized [18]. In the hospital admissions period, a median ensemble approach was utilized. Although included in figures as a benchmark, FluSight models were excluded from statistical analyses.

Forecasts were also classified as being generated from an ensemble of models or single models by either self-designation or review of their methods in the associated metadata. Ensembles were defined as those that combine the outputs of two or more forecasting models into a single submission, and single models referred to those utilizing only one model.

### Years of Participation

The years of participation variable refers to the number of seasons a team with a submitted model participated in the FluSight Challenge. This metric was used to assess whether greater participation in the Challenge was associated with better performance over time and across submitted models.

### Statistical Analysis

To assess the relationship between model type, years of participation, and performance, we conducted several statistical analyses. Spearman’s rank correlation was used to evaluate the relationship between years of participation and performance. Spearman’s rank correlation was chosen for its suitability with non-normally distributed and small sample data. Linear trend lines were generated using ordinary least squares regression to aid interpretation of relationships between participation history and forecast performance. Wilcoxon rank-sum tests were employed to compare performance between ensemble and single model forecasts and between ILI and hospital admissions seasons and hospital admissions seasons only participation groups. The Kruskal-Wallis test was applied to compare performance across multiple model types due to violations of normality assumptions in the performance data. Following significant results from the Kruskal-Wallis test, post-hoc pairwise comparisons were conducted using the Dunn’s test with p-values adjusted for multiple comparisons using the Holm method.

For the statistical tests, significance was set at an alpha level of 0.05. All analyses were conducted separately for the ILI and hospital admission periods to account for the differences in scoring methods and target datasets. See Supplementary Methods for detailed descriptions of each season’s methodology.

### Disclaimer

The findings and conclusions in this report are those of the authors and do not necessarily represent the views of the Centers for Disease Control and Prevention.

## Data Availability

The forecast data for each model are available from the FluSight Forecast Hub GitHub repositories (https://github.com/cdcepi/Flusight-forecast-data; https://github.com/cdcepi/FluSight-forecast-hub; https://github.com/cdcepi/FluSight-forecasts). These are all publicly accessible. The target data are also available as weekly counts for each jurisdiction from HHS.

## Acknowledgments

The authors would like to acknowledge Craig McGowan, Pragati Prasad, Jo Walker, Alexander Webber, Elizabeth White, and the following contributing institutions and teams.

Areté (ARETE), BioFire Diagnostics (BioFire), Catholic University of America (PPFST), Centre for Tropical Medicine and Global Health (FORSEA), Computational Epidemiology and Public Health Lab at Indiana University (CEPH), California Department of Public Health (CADPH), Carnegie Mellon University Delphi Group (Delphi, CMU), Center for Forecasting and Outbreak Analytics (CFA/CDC) (CFA_Pyrenew, cfa, cfarenewal), Centers for Disease Control and Prevention (FluSight), Columbia University (CU), Fjordhest (fjordhest), Fogarty International Center National Institutes of Health (NIH-Flu), Georgia Institute of Technology (Georgia Tech, Gatech), Georgia Institute of Technology Aditya Lab (GT), Georgia State University (Team_GA), Global Health Research Institute (GHRI), Google (Google), Google Science AI (Google_SAI), Guidehouse (GH Team A, GH Team B, Guidehouse, GH), Harvard University (02115 team, Harvard, ARGO), IBM Australia (IBM Australia), IEM Health (IEM Health), Indiana University (IU), Institute of Cognitive Science (Institute of Cognitive Science), Iowa State University (ISU), Iowa State University NiemiLab (ISU_NiemiLab), Jet Propulsion Laboratory California Institute of Technology (JPL), Johns Hopkins Infectious Disease Dynamics (JHU), Johns Hopkins University Applied Physics Laboratory (JHUAPL, APL), Kaiser Permanente Washington Health Research Institute (KPWHRI), Knowledge Based Systems Inc (KBSI), Korea Institute of Science and Technology Information (KIST), Lehigh University Computational Uncertainty Lab (LUcompUncertLab), Los Alamos National Laboratory (LANL), Los Alamos National Laboratory and Northern Arizona University (LosAlamos_NAU), Machine Intelligence Group for the betterment of Health and the Environment (MIGHTE), MOBS Lab at Northeastern University (MOBS), MOBS Lab at Northeastern University and ISI Foundation (NEU_ISI), Maryland Predict (MDPredict), Metaculus (Metaculus), Northeastern University (NEU, NEU-GLEAM), Northeastern University & University of California San Diego (NU_UCSD), One Health Trust and Johns Hopkins University (OHT_JHU), Predictive Science Inc. (PSI, PredSci, PSI-PROF), Protea Analytics (Protea Analytics), Rice University (Rice University), Signature Science (SigSci), Stevens Institute of Technology (Stevens), The Center for Systems Science and Engineering at Johns Hopkins University (JHU_CSSE), Tulane University (Tulane), UGA Center for the Ecology of Infectious Diseases (UGA_CEID), UNC Infectious Disease Dynamics (UNC_IDD), University of Arizona (UA, UA EpiFlu, UA-EpiCos, JL), University of California San Francisco (UCSF), University of Georgia (UGA, UGA_flucast), University of Guelph Dynamics Training Lab (UoGuelph, Uguelph, Uguelphensemble), University of Iowa (UI_CompEpi), University of Louisiana at Lafayette (ULL), University of Massachusetts Amherst (UMass, UMASS, KOT), University of Melbourne (uom, UniMelb), University of Michigan Computer Science and Engineering (UM), University of Minnesota (UMN, HumNat, UMNSpl), University of North Carolina (UNC), University of Southern California Srivastava Group (Sgroup), University of Texas at Austin (UT), University of Virginia (FluX, UVAFluX), University of Virginia, Biocomplexity Institute (UVA), Virginia Tech (4Sight, EpiDeep), Virginia Tech Sanghani Center for Artificial Intelligence and Data Analytics (VTSanghani), West Virginia Department of Health and Human Services (West Virginia DHHR), Yale University (Yale)

## Contributions

A.H., M.B., and R.K.B. contributed to conceptualization. A.H. and R.K.B. wrote the original draft of the manuscript. A.H. and S.M. performed data curation and formal analysis. M.B. and C.R. performed supervision and project administration. All authors contributed to the review and editing of the manuscript.

## References

1. Fischer, L.S., et al., CDC Grand Rounds: Modeling and Public Health Decision-Making. Morbidity and Mortality Weekly Report, 2016. 65(48): p. 1374–1377.

2. Biggerstaff, M., et al., Results from the centers for disease control and prevention’s predict the 2013-2014 Influenza Season Challenge. BMC Infect Dis, 2016. 16: p. 357.

3. Mathis, S.M., et al., Evaluation of FluSight influenza forecasting in the 2021-22 and 2022-23 seasons with a new target laboratory-confirmed influenza hospitalizations. medRxiv, 2023.

4. Borchering, R.K., et al., Responding to the Return of Influenza in the United States by Applying Centers for Disease Control and Prevention Surveillance, Analysis, and Modeling to Inform Understanding of Seasonal Influenza. JMIR Public Health Surveill, 2024. 10: p. e54340.

5. Reich, N.G., et al., Collaborative Hubs: Making the Most of Predictive Epidemic Modeling. Am J Public Health, 2022. 112(6): p. 839–842.

6. US COVID-19 Forecast Hub Consortium. The COVID-19 Forecast Hub. [cited 2026; Available from: https://covid19forecasthub.org/.

7. European Centre for Disease Prevention and Control. Respicast ECDC Respiratory Diseases Forecasting Hub. 2026; Available from: https://respicast.ecdc.europa.eu/.

8. Loo, S.L., et al., The US COVID-19 and Influenza Scenario Modeling Hubs: Delivering long-term projections to guide policy. Epidemics, 2024. 46: p. 100738.

9. Biggerstaff, M., et al., Results from the second year of a collaborative effort to forecast influenza seasons in the United States. Epidemics, 2018. 24: p. 26–33.

10. McGowan, C.J., et al., Collaborative efforts to forecast seasonal influenza in the United States, 2015-2016. Sci Rep, 2019. 9(1): p. 683.

11. U.S. Department of Health and Human Services COVID-19 Reported Patient Impact and Hospital Capacity by State Timeseries (RAW). 2023; Available from: https://healthdata.gov/Hospital/COVID-19-Reported-Patient-Impact-and-Hospital-Capa/g62h-syeh.

12. Shaman, J. and A. Karspeck, Forecasting seasonal outbreaks of influenza. Proc Natl Acad Sci U S A, 2012. 109(50): p. 20425–30.

13. Lutz, C.S., et al., Applying infectious disease forecasting to public health: a path forward using influenza forecasting examples. BMC Public Health, 2019. 19(1): p. 1659.

14. Bracher, J., et al., Evaluating epidemic forecasts in an interval format. PLoS Comput Biol, 2021. 17(2): p. e1008618.

15. Centers for Disease Control and Prevention. FluView Dashboard. 2025 April 11; Available from: https://gis.cdc.gov/grasp/fluview/fluportaldashboard.html.

16. Past Flu Season Severity Assessments. 2024; Available from: https://www.cdc.gov/flu/php/surveillance/past-seasons.html.

17. Centers for Disease Control and Prevention. Weekly Hospital Respiratory Data (HRD) Metrics by Jurisdiction. 2025; Available from: https://data.cdc.gov/Public-Health-Surveillance/Weekly-Hospital-Respiratory-Data-HRD-Metrics-by-Ju/mpgq-jmmr/about_data.

18. Reich, N.G., et al., Accuracy of real-time multi-model ensemble forecasts for seasonal influenza in the U.S. PLoS Comput Biol, 2019. 15(11): p. e1007486.

19. Centers for Disease Control and Prevention. FluSight Forecast Hub. 2025 April 21; Available from: https://github.com/cdcepi/FluSight-forecast-hub.

20. Johansson, M.A., et al., An open challenge to advance probabilistic forecasting for dengue epidemics. Proc Natl Acad Sci U S A, 2019. 116(48): p. 24268–24274.

21. Biggerstaff, M., et al., Improving Pandemic Response: Employing Mathematical Modeling to Confront Coronavirus Disease 2019. Clin Infect Dis, 2022. 74(5): p. 913–917.

22. Cramer, E.Y., et al., Evaluation of individual and ensemble probabilistic forecasts of COVID-19 mortality in the United States. Proc Natl Acad Sci U S A, 2022. 119(15): p. e2113561119.

23. U.S. Influenza Surveillance: Purpose and Methods. 2025; Available from: https://www.cdc.gov/fluview/overview/index.html.

24. Kandula, S., et al., Evaluation of mechanistic and statistical methods in forecasting influenza-like illness. J R Soc Interface, 2018. 15(144).

25. Ray, E.L., et al., Flusion: Integrating multiple data sources for accurate influenza predictions. Epidemics, 2025. 50: p. 100810.

26. Lopez, V.K., et al., Challenges of COVID-19 Case Forecasting in the US, 2020–2021. PLoS Comput Biol, 2024. 20(5): p. e1011200.

27. Bosse, N.I., et al., Scoring epidemiological forecasts on transformed scales. PLoS Comput Biol, 2023. 19(8): p. e1011393.

28. Centers for Disease Control and Prevention. FluSight 2024–2025 Evaluation. 2025; Available from: https://www.cdc.gov/flu-forecasting/evaluation/2024-2025-report.html.

29. Centers for Disease Control and Prevention. FluSight 2023–2024 Evaluation. 2025; Available from: https://www.cdc.gov/flu-forecasting/evaluation/2023-2024-report.html.

