## Supplementary Information for "A Decade of CDC FluSight Influenza Forecasting"

**Supplementary Information: A Decade of the Centers for Disease Control and Prevention's FluSight Influenza Forecasting**

**Supplementary Table 1. United States Influenza Season Details 2014/15 - 2024/25**

Seasonal epidemiologic characteristics used to contextualize forecasting performance, including predominant influenza virus strain, peak month, peak ILI intensity, CDC severity classification, national baseline ILI level, and duration of elevated activity measured as weeks above baseline[1, 2].

| | Predominant Virus* | Season Peak | Peak Intensity (% ILI) | Overall Severity [3] | National Baseline** (ILI%) | Length (weeks above baseline) |
|---|---|---|---|---|---|---|
| 2014/15 | A/H3N2 | December | 6 | High | 2 | 19 |
| 2015/16 | A/H1N1pdm09 | March | 3.6 | Moderate | 2.1 | 12 |
| 2016/17 | A/H3N2 | February | 5.1 | Moderate | 2.2 | 16 |
| 2017/18 | A/H3N2 | February | 7.5 | High | 2.2 | 19 |
| 2018/19 | A/H1N1pdm09, A/H3N2 | February | 5 | Moderate | 2.2 | 20 |
| 2019/20 | A/H1N1pdm09, B | December | 7.1 | Moderate | 2.4 | 23 |
| 2020/21 | Minimal Influenza Activity | NA | 2.3 | Low | 2.6 | 0 (never exceeded national baseline) |
| 2021/22 | A/H3N2 | December | 4.9 | Low | 2.5 | 9 |
| 2022/23 | A/H3N2 | November | 7.4 | Moderate | 2.5 | 21 |
| 2023/24 | A/H1N1pdm09 | December | 6.8 | Moderate | 2.9 | 22 |
| 2024/25 | A/H1N1pdm09, A/H3N2 | February | 7.8 | Severe [4] | 3 | 17 |

* The virus that made up the largest proportion of all influenza tests reported to CDC by public health laboratories.

** Baseline threshold is developed by calculating the mean percentage of patient visits for ILI during non-influenza weeks for the most recent two seasons and adding two standard deviations. A non-influenza time period (e.g., a "non-influenza week") is defined as two or more consecutive weeks in which each week accounted for less than 2% of the season's total number of specimens that tested positive for influenza in public health laboratories [5].

**Supplementary Table 2. Model Type vs Performance Results by Season (screenshot below table in case of formatting issue)**

Kruskal-Wallis and post hoc Dunn test results comparing model performance by type across influenza seasons. The table includes the overall Kruskal-Wallis test results for each season and from the hospital admissions seasons as a group, followed by pairwise comparisons among model types using Dunn's test with Holm adjustment. Post hoc comparisons were only conducted for seasons with a statistically significant Kruskal-Wallis result. NA indicates that the Dunn test was not performed. Asterisk (*)

indicates a statistically significant difference at the 0.05 level (adjusted p-value). The model type listed first is the better performing type in the bolded comparisons. These pooled results should be interpreted with caution because scoring criteria varied across ILI seasons. Model types included: mechanistic models (Mech), statistical models (Stat), machine learning models (ML), and statistical/mechanistic hybrid models (Stat/mech) (See methods).

| Season | Kruskal-Wallis H (p-value) | Comparison | Dunn Adjusted p-value |
|---|---|---|---|
| 2014/15 | 1.125 (0.2888) | NA | NA |
| 2015/16 | 1.667 (0.1967) | NA | NA |
| 2016/17 | 7.995 (0.0461) * | Stat/mech vs. Mech | 0.0885 |
| | | Stat vs. ML | 0.3362 |
| | | Stat vs. Mech | 0.5030 |
| | | Mech vs. ML | 0.5968 |
| | | Stat/mech vs. ML | 0.1444 |
| | | Stat vs. Stat/mech | 0.4529 |
| 2017/18 | 1.3315 (0.7217) | NA | NA |
| 2018/19 | 8.838 (0.0315)* | **Stat vs. Mech** | **0.0323*** |
| | | Stat vs. ML | 0.2202 |
| | | Mech vs. ML | 1.000 |
| | | Stat/mech vs. ML | 0.7016 |
| | | Stat vs. Stat/mech | 1.000 |
| | | Stat/mech vs. Mech | 0.9799 |
| 2019/20 | 11.19 (0.0107)* | **Stat vs. ML** | **0.0273*** |
| | | Stat vs. Mech | 0.2412 |
| | | Stat/mech vs. ML | 0.1028 |
| | | Stat/mech vs. Mech | 0.2294 |
| | | Mech vs. ML | 0.8169 |
| | | Stat vs. Stat/mech | 1.000 |
| ILI Period (2014/15-2019/20) | 16.157 (0.001)* | **Stat vs. Mech** | **0.0041*** |
| | | **Stat/mech vs. Mech** | **0.0335*** |
| | | Stat vs. ML | 0.0621 |
| | | Stat/mech vs. ML | 0.0964 |
| | | Mech vs. ML | 1.000 |
| | | Stat vs. Stat/mech | 0.6571 |
| 2021/22 | 3.345 (0.3415) | NA | NA |
| 2022/23 | 2.573 (0.4622) | NA | NA |
| 2023/24 | 3.716 (0.2938) | NA | NA |
| 2024/25 | 5.327 (0.1494) | NA | NA |
| Hospital Admissions Period (2021/22-2023/24) | 8.920 (0.0922)* | NA | NA |

### Supplementary Table 3. Top 5 Models in each Season of the FluSight Challenge from 2015/16 - 2023/24

Teams from the 2013/14 and 2014/15 seasons were not included in this table because the participants were kept anonymous, with the exception of the winning teams. The designated (publicly communicated) FluSight ensemble for 2015/16 through 2017/18 was an unweighted mean ensemble (FluSight-Ensemble). A weighted ensemble, with forecast weights based on previous forecast model performance was piloted in 2017/18 and became the designated ensemble in 2018/19 (FluSight-Network). In the hospital admissions seasons (2021/22 to 2024/25), the designated FluSight ensemble was an unweighted median ensemble (FluSight-ensemble).

| Season | 1st | 2nd | 3rd | 4th | 5th |
|---|---|---|---|---|---|
| 2015/16 | **FluSight-Ensemble (ENS)** | Delphi-Archefilter (STAT) | Delphi-Epicast (STAT, ENS) | Kernel of Truth (STAT) | Delphi-Stat (STAT, ENS) |
| 2016/17 | Delphi-Epicast (STAT, ENS) | CU3 (STAT, ENS) | CU1 (STAT, ENS) | Delphi-Stat (STAT, ENS) | CU2 (STAT/MECH, ENS) |
| 2017/18 | Delphi-Epicast (STAT, ENS) | **FluSight-Network (ENS)*** | LANL-DBMplus (STAT/MECH) | LANL-DBM (STAT/MECH) | CU_Vixen (MECH, ENS) |
| 2018/19 | LANL-Dante (STAT) | LANL-DBMplus (STAT/MECH) | **FluSight-Network (ENS)** | PPFST (STAT, ENS) | PPFST (STAT, ENS) |
| 2019/20 | LANL-DBMplus (STAT/MECH) | **FluSight-Network (ENS)** | CU_Ravens (STAT, ENS) | Protea-Springbok (STAT) | Protea-Cheetah (STAT, ENS) |
| 2021/22 [6] | CMU-TimeSeries (STAT) | **FluSight-ensemble (ENS)** | PSI-DICE (MECH) | UMass-trends_ensemble (STAT, ENS) | SGroup-RandomForest (ML, ENS) |
| 2022/23 [6] | MOBS-GLEAM_FLUH (MECH) | CMU-TimeSeries (STAT, ENS) | PSI-DICE (MECH) | MIGHTE-Nsemble (ML, ENS) | **FluSight-ensemble (ENS)** |
| 2023/24 [7] | UMass-flusion (ML, ENS) | **FluSight-ensemble (ENS)** | MIGHTE-Nsemble (ML, ENS) | UGA_flucast-INFLAenza (ML) | CU-ensemble (ENS, MECH, AI/ML, STAT) |
| 2024/25 [8] | **FluSight-ensemble (ENS)** | PSI-PROF_beta (STAT/MECH) | CU-ensemble (ENS, MECH, AI/ML, STAT) | NEU_ISI-AdaptiveEnsemble (STAT, ENS) | CMU-TimeSeries (ENS, STAT) |

*FluSight-Network ensemble was piloted in 2017/18 season, but it was not designated as the publicly communicated FluSight ensemble until the 2018/19 season [9].

**Supplementary Fig. 1. Boxplots comparing model type performance in pooled ILI and hospital admissions influenza seasons.**

A. Influenza-like Illness seasons (2014/15-2019/20). Distribution of skill for each model type; higher values indicate better performance. These pooled results should be interpreted with caution because scoring criteria varied across ILI seasons.
B. Hospital admissions seasons (2021/22-2024/25). Distribution of Relative Weighted Interval Score (WIS) for each model type; lower values indicate better performance. Relative WIS is displayed on a logarithmic scale with baseline performance equal to one.

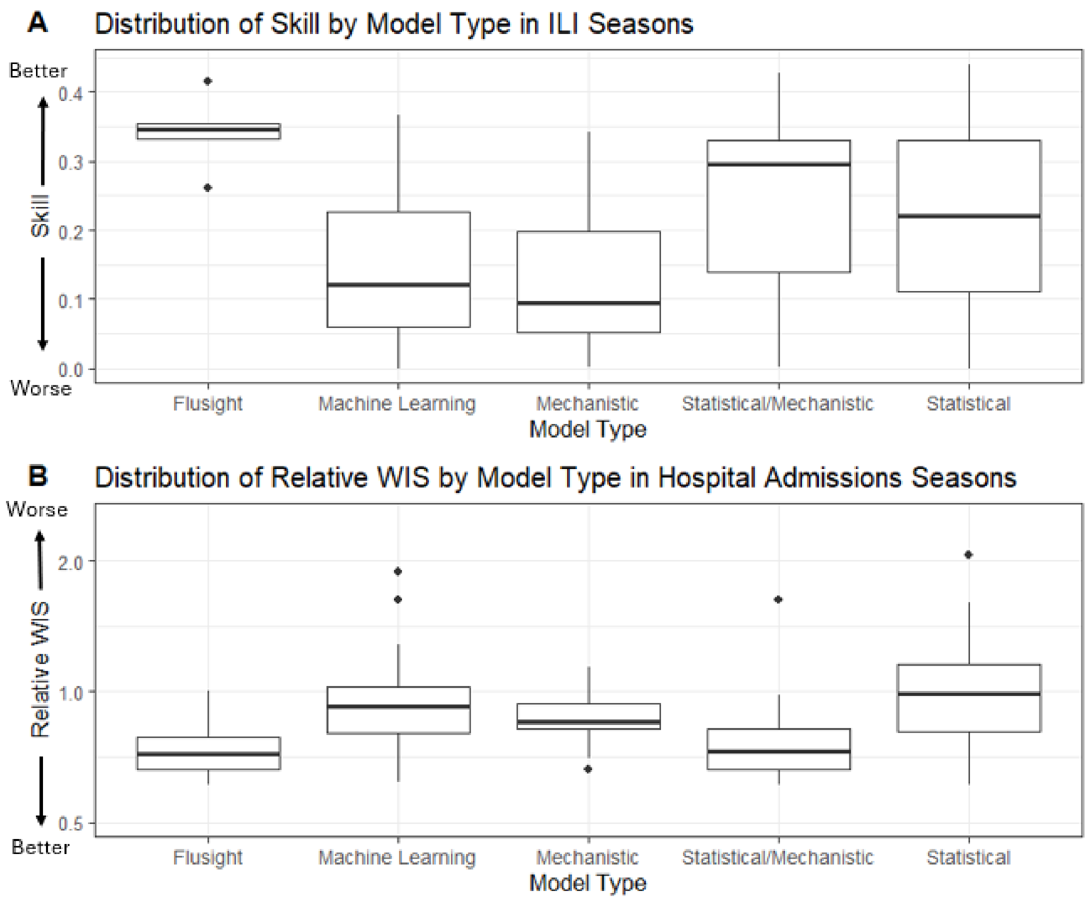


**Supplementary Fig. 2. Boxplots comparing submitted ensemble and single model performance in pooled ILI and hospital admissions influenza seasons.**

A. Influenza-like Illness seasons (2014/15-2019/20). Distribution of skill by ensemble or single model status; higher values indicate better performance. These pooled results should be interpreted with caution because of scoring criteria varied across ILI seasons.
B. Hospital admissions seasons (2021/22-2024/25). Distribution of relative Weighted Interval Scores (WIS) by ensemble or single model status; lower values indicate better performance. Relative WIS is displayed on a logarithmic scale with baseline performance equal to one.
Asterisks (*) denote statistically significant differences between groups ($p < 0.05$).

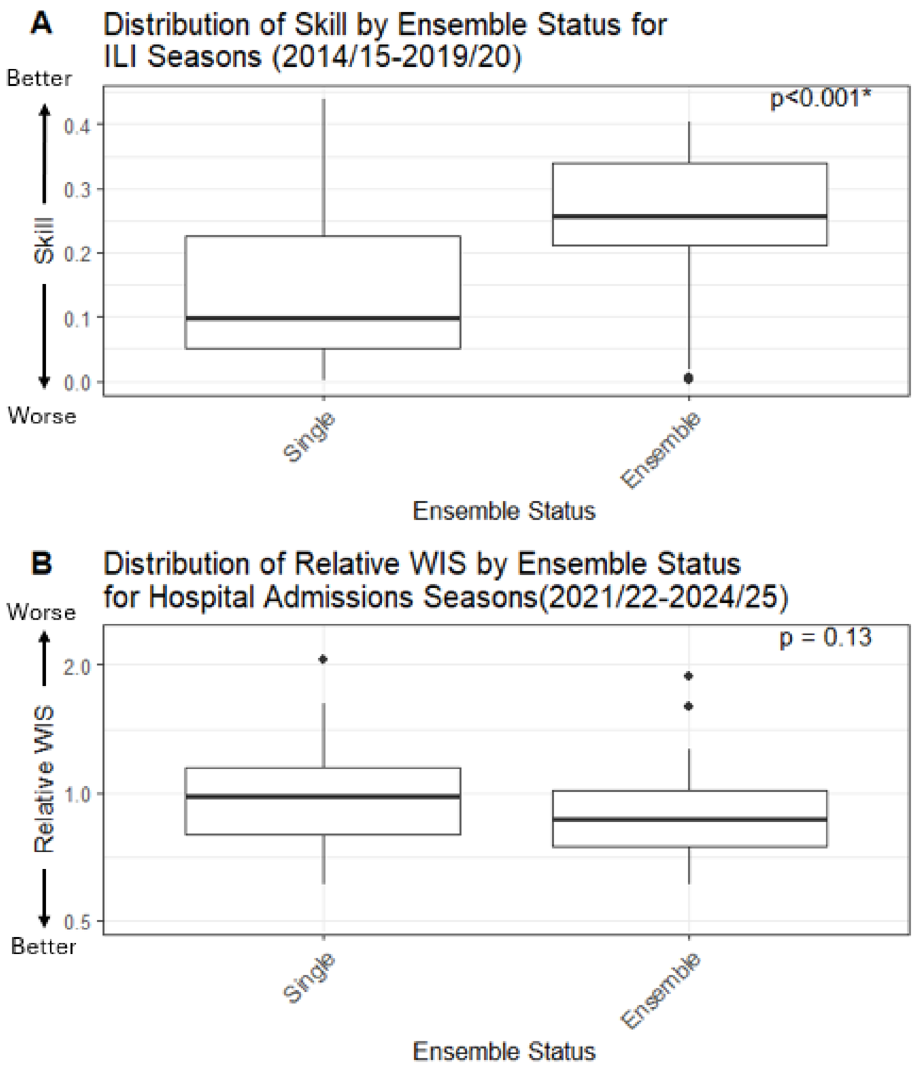


**Supplementary Fig. 3. Scatter plots show the relationship between model performance and number of participation years for pooled ILI and hospital admissions influenza seasons.**

A. Influenza-like Illness seasons (2014/15-2019/20): Skill plotted against participation years, with trend line and correlation coefficient. Higher skill indicates better performance. These pooled results should be interpreted with caution because of scoring criteria varied across ILI seasons.
B. Hospital admissions seasons (2021/22-2024/25): Relative Weighted Interval Score (WIS) plotted against participation years, with trend line and correlation coefficient. Lower Relative WIS values indicate better performance. Relative WIS is displayed on a logarithmic scale with baseline performance equal to one.
Asterisks (*) indicate statistically significant correlations ($p < 0.05$).

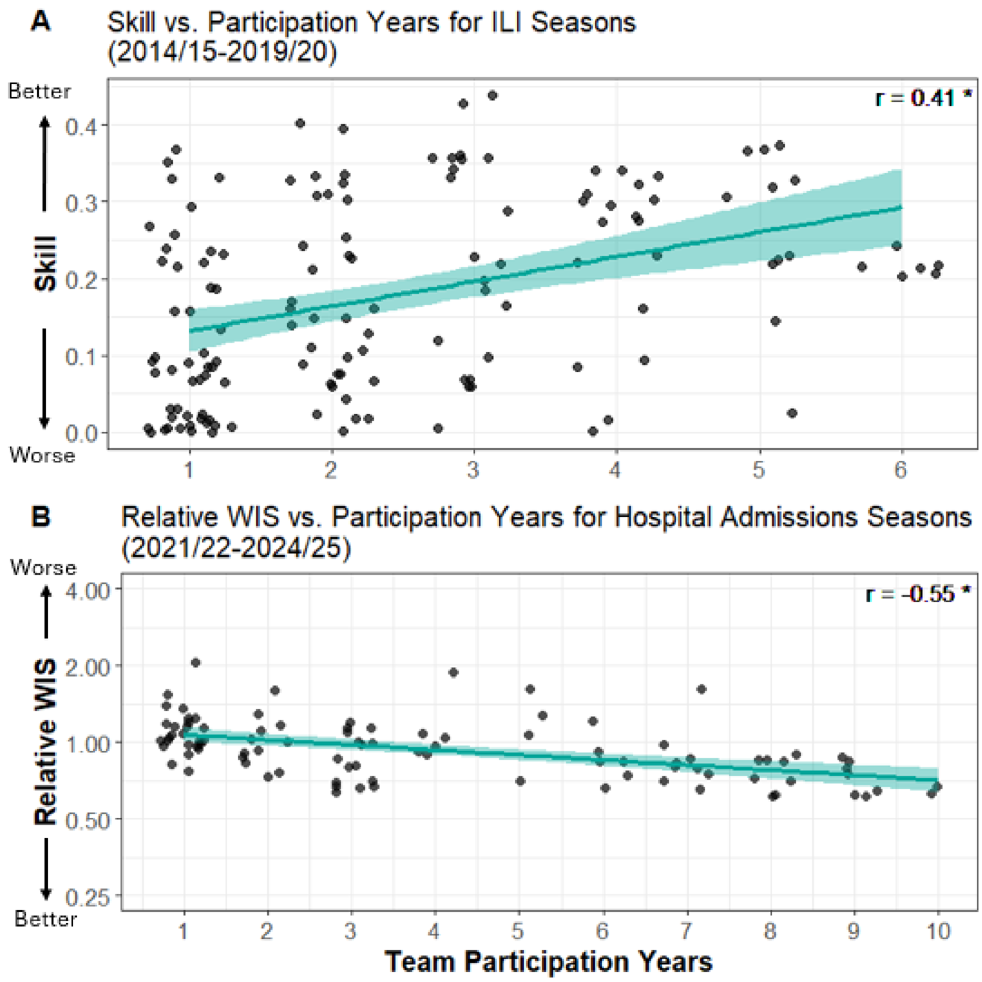


**<u>Supplementary Methods: Comprehensive Season Scoring Criteria 2013/14 - 2024/25</u>**

**2013/14**:

The 2013/14 FluSight Challenge ran from December 1, 2013, to March 27, 2014, during which teams submitted nine biweekly forecasts for national-level seasonal influenza milestones: onset, peak week, duration, and intensity[10]. To be eligible for judging, teams were required to incorporate at least one form of digital surveillance data (e.g., Twitter, internet search trends, or online surveys), though integration with traditional surveillance data such as ILINet was permitted. Submissions also required an accompanying narrative detailing the model methodology, which could be updated throughout the contest to reflect methodological changes.

All forecasts were evaluated against ILINet data. The start of the season was defined as the first week in which the weighted ILI percentage exceeded the national baseline of 2.0% and remained above it for at least two additional weeks. The peak week corresponded to the week with the highest ILI percentage, and peak intensity to the maximum observed ILI percentage. Season duration was defined as the number of weeks ILI remained above the baseline.

Accurate forecasts were defined as those which were within one week of the observed value (for timing targets) or within one percentage point (for intensity targets) of the observed values. Teams could also submit analagous forecasts for any or all the 10 HHS regions, which contributed to their final scores as described below.

Submissions were evaluated by a panel of three judges—two from CDC and one academic reviewer—using a 100-point scale: methodological strength (25 points), including clarity and uncertainty communication; forecast accuracy, timeliness, and reliability (65 points); and geographic scope (10 points). Up to 50 bonus points were awarded to any contestant that submitted forecasts for the 10 HHS regions; the number of bonus points was based on the number of regions with a forecast and the strength of the methodology, and the accuracy, timeliness, and reliability of the HHS region forecasts.

**2014/15:**

The 2014/15 FluSight Challenge required weekly forecasts from October 20, 2014, through May 25, 2015[11]. Forecast targets included three seasonal outcomes—onset week, peak week, and peak intensity—as well as four short-term forecasts of wILI% (ILI percentage weighted by state population from ILINet) one to four weeks ahead. Forecasts were submitted at the national and HHS regional levels. Definitions of the seasonal targets remained consistent with the previous season.

Forecasts consisted of both point estimates and probabilistic distributions represented in predefined bins. For onset and peak week, bins corresponded to individual MMWR weeks, with an additional bin for "no onset." For peak intensity and short-term forecasts, bins used semi-open intervals of 1% width, ranging from 0% to 10%, with an additional bin for values exceeding 10%.

Forecast accuracy was evaluated using the logarithmic scoring rule. A forecast's log score was defined as $S(\mathbf{p},i)=\ln(p_i)$, where $p_i$ is the probability assigned to the observed outcome. Forecasts assigning zero probability to the true outcome or failing to define a valid probability distribution were assigned a default score of -10. Scores were averaged across all locations, targets, and weeks. For interpretability, mean log scores were exponentiated to yield a "forecast skill" metric ranging from 0 to 1.

**2015/16:**

The 2015/16 FluSight Challenge largely followed the format of the previous season, with weekly submissions from November 2, 2015, to May 16, 2016[12]. Forecasting targets remained unchanged: onset week, peak week, and peak intensity, as well as short-term predictions of wILI% one to four weeks ahead. Teams submitted both point estimates and probabilistic forecasts at national and regional levels.

A key change in the 2015/2016 season was the refinement of the binning structure for intensity and short-term forecasts, which used 0.5% intervals ranging from 0% to 12.5%, with an additional bin for values equal to or over 13%. The log scoring rule remained in place, with exponentiated mean log scores used to calculate forecast skill. Forecasts that are missed, incomplete, or when the probability assigned to the observed outcome is 0 were assigned a score of -10.

As in prior seasons, participants submitted methodological narratives, which could be updated during the season. Two benchmark models were introduced: a historical average model based on kernel density estimation from past seasons of observed ILI data and an unweighted ensemble forecast (FluSight Ensemble), which averaged the predictive distributions submitted by all teams each week.

All forecasts were scored using finalized ILINet data published in MMWR week 28, with evaluation considering the bin containing the observed outcome and the immediately adjacent bins as accurate.

**2016/17:**

The 2016/17 FluSight Challenge retained the core structure of previous seasons. Participants submitted weekly forecasts of ILI activity from November 7, 2016, through May 15, 2017[13]. Forecasts were required at the national level and encouraged for the 10 HHS regions. Each submission included point predictions and full probabilistic distributions for seasonal targets (onset week, peak week, and peak intensity) and short-term targets (wILI% one to four weeks ahead). All forecasts were evaluated against ILINet data.

Target definitions remained consistent with prior seasons. Onset was defined as the first of three consecutive MMWR weeks in which wILI% met or exceeded region-specific baseline values. Peak week referred to the MMWR week with the highest observed wILI%, and peak intensity was the highest rounded wILI% observed in the season. In the event of ties for peak week, all tied weeks were considered valid. ILINet percentages were rounded to one decimal place before scoring.

Probabilistic forecasts were submitted in predefined bins: 1-week intervals for temporal targets, and 0.1% intervals for intensity and short-term wILI% targets, ranging from 0% to 12.9%, with an additional bin for values ≥13%. For onset, teams could include a “no onset” category if they anticipated that the baseline thresholds would not be met. Forecasts were required to sum to 1.0 (or fall within a normalization range of 0.9–1.1), and invalid or improperly normalized submissions were penalized.

Forecasts were scored using a logarithmic scoring rule. For onset and peak week, the log score included the bin corresponding to the observed outcome as well as the bins immediately before and after. For intensity and short-term forecasts, the score was based on the observed bin and the five adjacent bins on either side. Any forecast assigning zero probability to the observed outcome or submitting an invalid distribution was assigned a score of -10. Final forecast skill was calculated by averaging log scores across time points, locations, and targets, and then exponentiating to improve interpretability.

This season continued to utilize the historic baseline model and unweighted ensemble as described in the 2015/16 season. Teams could elect whether their forecasts would be included in the ensemble and how they would be attributed (public, anonymous, or withheld).

While the primary evaluation was based on log scores of probabilistic forecasts, point predictions were collected for secondary analyses of absolute error.

**2017/18:**

The 2017/18 FluSight Challenge continued the weekly forecasting structure, requiring submissions from November 6, 2017, through May 14, 2018[13]. Participants submitted required national-level forecasts, with regional forecasts encouraged. Forecasts could include any or all of the seasonal targets (onset week, peak week, peak intensity) and short-term targets (wILI% one to four weeks ahead), using both point predictions and full predictive distributions. All forecasts were evaluated against ILINet data. Target definitions remained unchanged and ILINet values were rounded to one decimal place for evaluation.

Forecasts were required to follow a standardized CSV format with fixed structure. Probabilities had to be assigned across predefined bins that summed to 1.0 (or within a 0.9–1.1 normalization range). Forecasts outside this range were normalized or discarded, and missing or invalid submissions were penalized. For onset, teams could assign probability to a "no onset" bin for seasons in which the baseline threshold was not expected to be crossed.

Scoring was based on the logarithmic scoring rule, consistent with earlier seasons. For temporal targets (onset and peak week), the score considered the observed bin and the adjacent bins as accurate. For peak intensity and short-term targets, the score considered the observed bin and the five bins on either side as accurate. Any forecast that assigned zero probability to the observed outcome received a score of -10, as did missing or improperly formatted forecasts. While point forecasts were not part of the primary evaluation, they were analyzed separately using absolute error.

Final team rankings were based on average log scores across time, targets, and locations, during a defined evaluation period beginning four weeks before the observed onset and ending three weeks after wILI% fell below baseline. Participants could update their models throughout the season, provided they submitted revised narratives documenting methodological changes. Teams were encouraged to draw from external data sources such as those provided in CMU's Epidata API, HealthTweets, and historical ILINet data from the FluView portal or 'cdcfluview' R package.

**2018/19:**

The 2018/19 FluSight Challenge invited individuals and teams to submit weekly forecasts of influenza activity from October 29, 2018, through May 13, 2019[13]. Participants provided probabilistic and point forecasts for three seasonal targets, onset week, peak week, and peak intensity, and short-term targets of wILI% one to four weeks ahead. Forecasts were submitted at both the national and HHS regional levels, and all evaluations were based on ILINet data. Target definitions remained unchanged and ILINet values were rounded to one decimal place for evaluation.

Forecasts were submitted in a standardized CSV format and were required to include non-negative probability distributions that summed to 1.0 or within a normalization range of 0.9–1.1. Probabilities falling outside this range led to forecasts being discarded. For onset, probability could be assigned to a "no onset" option indicate a forecast of not exceeding the baseline threshold during the season. Probabilistic bins were defined at 1-week intervals for temporal targets and at 0.1% intervals for intensity and short-term targets, with an additional bin for values ≥13%.

All forecasts were evaluated using a logarithmic scoring rule. For onset and peak week, the score included the observed bin and the adjacent bins (±1 week). For intensity and short-term targets, the observed bin and the five bins on either side were considered accurate and contributed to the score. Forecasts assigning zero probability to the observed bin or failing to sum correctly were assigned a score of -10. Point forecasts were not used for ranking but were evaluated separately using absolute error.

Final scores were averaged across time, locations, and targets to determine team rankings. Evaluation periods varied by target type: seasonal targets were scored from the start of the season through six weeks after onset (onset week target) or until wILI% fell below baseline (peak week and intensity targets); short-term forecasts were evaluated from four weeks before observed onset through three weeks after the return below baseline.

As in prior seasons, participants were encouraged to submit methodological descriptions and could update them during the Challenge. Forecasts were benchmarked against a historical average model and a real-time ensemble developed by the FluSight Network, which incorporated past performance-based weighting. Participants could choose to contribute to this ensemble and select their level of attribution (public, anonymous, or withheld).

**2019/20:**

The 2019/20 Challenge season invited participants to submit weekly probabilistic forecasts for seven national and regional targets: three seasonal (onset week, peak week, and peak intensity) and four short-term (1- to 4-week ahead ILI values)[13]. The Challenge was planned to run from October 28, 2019, to May 11, 2020, with forecasts due each Monday by 11:59 PM ET. However, due to declines in influenza activity associated with the COVID-19 pandemic, the Challenge was declared over on March 10, 2020. Late or missed submissions were accepted a score of –10.

Season onset was defined as the first of three consecutive weeks where weighted ILI met or exceeded the regional baseline. Peak week was the week with the highest observed wILI%, and peak intensity was the maximum weighted wILI% during the season. Short-term forecasts corresponded to ILI percentages one to four weeks after the most recently published ILINet data. All forecasts were required at both the national level and for each of the 10 HHS regions.

Forecasts were probabilistic and submitted in a fixed CSV format. Each forecast included probabilities across defined bins and a point prediction. Probabilities had to be non-negative and sum to between 0.9 and 1.1; otherwise, forecasts were discarded and assigned a score of -10.

Forecast accuracy was evaluated using the logarithmic scoring rule, based on the probability assigned to the bin containing the observed value. Multiple peak weeks were scored by summing probabilities assigned to all correct bins. A probability of zero for the observed bin received a score of -10. Point predictions were used to compute absolute error, although only log scores contributed to final team rankings.

Teams were required to submit all seven targets for all locations at least once to be eligible for overall ranking. Final scores were computed as the average log score across all targets and jurisdictions over their evaluation windows. Evaluation periods varied by target type: seasonal targets were scored from the start of the season through six weeks after onset (for onset) or when wILI% dropped below baseline (for peak week and intensity); short-term forecasts were evaluated from four weeks before observed onset through three weeks after the return below baseline.

For short-term targets, evaluation ran from four weeks before onset to three weeks after wILI% returned below baseline.

Historical ILI data and baselines were provided via the CDC FluView portal and GitHub repositories. Utilizing additional data sources, including those in the Delphi Epidata API and social media signals, were permitted. All forecasts were published on the CDC Epidemic Prediction Initiative website and GitHub. Teams could choose to be identified or remain anonymous.

An ensemble forecast created by the FluSight Network was submitted weekly using past performance-based weighting. All forecasts were also compared to a historical model based on past ILI data.

**2021/22:**

The 2021/22 influenza season included a shift to weekly forecasts of confirmed influenza hospital admissions as the primary target data source[6]. Beginning January 10, 2022, participants submitted national- and state-level probabilistic forecasts for 1- to 4-week-ahead confirmed hospital admissions. Submissions were due each Monday by 11:00 PM ET and evaluated against the previous_day_admission_influenza_confirmed field (Field #34) from the COVID-19 Reported Patient Impact and Hospital Capacity by State Timeseries dataset, aggregated weekly using MMWR epidemiological week definitions.

Seasonal target forecasts (e.g., onset week or peak week) were not collected for this season. Data preprocessing included shifting reported admissions back one day to align with admission dates. National-level admissions data were derived by summing admissions across all U.S. states, D.C., Puerto Rico, and the U.S. Virgin Islands.

Forecasts were submitted in a standardized CSV format, containing both point and quantile forecasts across 23 quantiles: 0.01, 0.025, every 0.05 increment from 0.05 to 0.95, and then 0.975 and 0.99. Each submission file followed a strict naming and folder structure and included required metadata. Teams designated models as “primary,” “secondary,” “proposed,” or “other” to determine eligibility for ensemble inclusion and evaluation.

Forecast accuracy was evaluated using weighted interval score (WIS) and prediction interval coverage. All forecasts were published in real time on CDC’s GitHub repository and were included in ensemble models based on their designation or included in scientific publications summarizing results. Historical hospitalization data from the 2020–21 season were available for model fitting, and teams could use additional external data sources such as those provided in the CDC FluView portal and the Carnegie Mellon Epidata API.

**2022/23:**

The 2022/23 influenza hospital admissions forecasting Challenge ran from October 17, 2022, to May 22, 2023[6]. Participating teams submitted weekly national- and state-level probabilistic forecasts for the number of laboratory-confirmed influenza hospital admissions 1- to 4- weeks ahead. Forecasts were evaluated against the previous_day_admission_influenza_confirmed field (Field #34) from the COVID-19 Reported Patient Impact and Hospital Capacity by State Timeseries dataset, aggregated using CDC epidemiological weeks (Sunday–Saturday). Forecasts were submitted to the FluSight GitHub repository [14].

Seasonal target forecasts (e.g., peak week and onset week) were not collected in this season, although an experimental rate trend target (e.g., “likely increase/decrease”) was introduced mid-season. Forecasts adhered to a standardized CSV format and file structure, with teams submitting quantile and point forecasts across 23 quantiles.

Hospital admission data were adjusted by shifting admissions one day earlier to align with admission dates, and national totals included all 50 states, D.C., Puerto Rico, and the U.S. Virgin Islands. Data reporting compliance improved after February 2022 when Fields 33–35 became mandatory for hospitals.

Forecast accuracy was assessed using weighted interval score (WIS) and prediction interval coverage, with evaluations based on data as of six weeks after the final forecast due date. Submissions were hosted publicly via the CDC GitHub repository and could be included in ensemble models based on their designation in the associated metadata.

**2023/24:**

The 2023/24 FluSight Challenge ran from October 11, 2023, to May 1, 2024. Participating teams submitted weekly national- and state-level probabilistic forecasts for the number of laboratory-confirmed influenza hospital admissions. In this season, short-term targets were defined as 0- to 3-weeks ahead, reflecting a shift from the previous 1- to 4-week-ahead definition used in earlier seasons due to changes in data release timing. Forecasts were evaluated against the previous_day_admission_influenza_confirmed field (Field #34) from the COVID-19 Reported Patient Impact and Hospital Capacity by State Timeseries dataset, aggregated using CDC epidemiological weeks (Sunday–Saturday). Forecasts were submitted to the FluSight GitHub repository [14].

The experimental rate trend target was included as an optional target following the pilot of it in the 2022/23 season. Specific rate-difference thresholds for changes were developed for each prediction horizon, based on past distributions observed in FluSurv-NET and HHS-Protect data. Forecasts adhered to a standardized CSV format and file structure, with teams submitting point and probabilistic forecasts across 23 quantiles.

Hospital admission data were adjusted by shifting admissions one day earlier to align with admission dates, and national totals included all 50 states, D.C., Puerto Rico, and the U.S. Virgin Islands. Data reporting compliance improved after February 2022 when Fields 33–35 became mandatory for hospitals.

Forecast accuracy was assessed using weighted interval score (WIS) and prediction interval coverage, with evaluations based on data as of six weeks after the final forecast due date. Submissions were hosted publicly via the CDC GitHub repository and were included in the FluSight ensemble models based on their designation in the associated metadata.

**2024/25:**

The 2024/25 FluSight Challenge ran from November 20, 2024, to May 31, 2025. Participating teams submitted weekly national- and state-level probabilistic forecasts for the number of laboratory-confirmed influenza hospital admissions. In this season, short-term targets were defined as 0- to 3-weeks ahead. Forecasts were evaluated against the 'total number of new hospital admissions of patients with confirmed influenza captured during the reporting week' field from the Weekly Hospital Respiratory Data (HRD)

Metrics by Jurisdiction, National Healthcare Safety Network (NHSN) [15]. Forecasts were submitted to the FluSight GitHub repository [14].

For the rate trend target, rate-difference thresholds were updated for each prediction horizon based on past distributions observed in FluSurv-NET and HHS-Protect data [6]. Seasonal targets including peak week and peak incidence for the season were included as new optional targets. Forecasts were submitted in a standard CSV format and file structure, with teams submitting probabilistic forecasts across 23 quantiles.

Forecast accuracy was assessed using weighted interval score (WIS) and prediction interval coverage, with evaluations based on data as of six weeks after the final forecast due date [8]. Submissions were hosted publicly via the CDC GitHub repository and were included in the FluSight ensemble models based on their designation in the associated metadata.

### References


1. Centers for Disease Control and Prevention. *FluView Dashboard*. 2025 April 11; Available from: https://gis.cdc.gov/grasp/fluview/fluportaldashboard.html.
2. Centers for Disease Control and Prevention. *About Influenza*. 2026 [cited 2026 May 5]; Available from: https://www.cdc.gov/flu/about/index.html#cdc_disease_basics_quick_facts_callout_callout-quick-facts.
3. Centers for Disease Control and Prevention. *Past Flu Season Severity Assessments*. 2024; Available from: https://www.cdc.gov/flu/php/surveillance/past-seasons.html.
4. Centers for Disease Control and Prevention. *Influenza Activity in the United States during the 2024–25 Season and Composition of the 2025–26 Influenza Vaccine*. 2025; Available from: https://www.cdc.gov/flu/whats-new/2025-2026-influenza-activity.html.
5. Centers for Disease Control and Prevention. *U.S. Influenza Surveillance: Purpose and Methods*. 2025; Available from: https://www.cdc.gov/fluview/overview/.
6. Mathis, S.M., et al., *Evaluation of FluSight influenza forecasting in the 2021-22 and 2022-23 seasons with a new target laboratory-confirmed influenza hospitalizations.* medRxiv, 2023.
7. Centers for Disease Control and Prevention. *FluSight 2023-2024 Evaluation*. 2025; Available from: https://www.cdc.gov/flu-forecasting/evaluation/2023-2024-report.html.
8. Centers for Disease Control and Prevention. *FluSight 2024-2025 Evaluation*. 2025; Available from: https://www.cdc.gov/flu-forecasting/evaluation/2024-2025-report.html.
9. Reich, N.G., et al., *Accuracy of real-time multi-model ensemble forecasts for seasonal influenza in the U.S.* PLoS Comput Biol, 2019. **15**(11): p. e1007486.
10. Biggerstaff, M., et al., *Results from the centers for disease control and prevention's predict the 2013-2014 Influenza Season Challenge.* BMC Infect Dis, 2016. **16**: p. 357.
11. Biggerstaff, M., et al., *Results from the second year of a collaborative effort to forecast influenza seasons in the United States.* Epidemics, 2018. **24**: p. 26-33.
12. McGowan, C.J., et al., *Collaborative efforts to forecast seasonal influenza in the United States, 2015-2016.* Sci Rep, 2019. **9**(1): p. 683.
13. Centers for Disease Control and Prevention. *FluSight Forecasts*. 2024; Available from: https://github.com/cdcepi/FluSight-forecasts.

14. Centers for Disease Control and Prevention. *FluSight Forecast Hub*. 2025 April 21; Available from: https://github.com/cdcepi/FluSight-forecast-hub.
15. Centers for Disease Control and Prevention. *Weekly Hospital Respiratory Data (HRD) Metrics by Jurisdiction*. 2025; Available from: https://data.cdc.gov/Public-Health-Surveillance/Weekly-Hospital-Respiratory-Data-HRD-Metrics-by-Ju/mpgq-jmmr/about_data.
